# Can a greenhouse gas emissions tax on food also be healthy and equitable? A systematized review and modelling study from Aotearoa New Zealand

**DOI:** 10.1101/2022.02.15.22271015

**Authors:** Christine Cleghorn, Ingrid Mulder, Alex Macmillan, Anja Mizdrak, Jonathan Drew, Nhung Nghiem, Tony Blakely, Cliona Ni Mhurchu

**Affiliations:** Burden of Disease Epidemiology, Equity and Cost-Effectiveness Programme, University of Otago, Wellington, New Zealand; Department of Preventive and Social Medicine, University of Otago, Dunedin, Aotearoa New Zealand; Population Interventions, Centre for Epidemiology and Biostatistics, Melbourne School of Population and Global Health, University of Melbourne, Melbourne, Australia; National Institute for Health Innovation, University of Auckland, Private Bag 92019, Auckland Mail Centre, Auckland 1142, New Zealand; The George Institute for Global Health, 1 King Street, Newtown 2042 New South Wales, Australia

## Abstract

**Introduction:** Policies to mitigate climate change are essential. The objective of this paper was to estimate the impact of greenhouse gas (GHG) food taxes and assess whether such a tax could also have health benefits and reduce ethnic inequalities in health in Aotearoa NZ.

**Methods:** We undertook a systemised review on GHG food taxes to inform four tax scenarios, including one combined with a subsidy. These scenarios were modelled to estimate lifetime impacts on quality adjusted health years (QALY), health inequities by ethnicity, GHG emissions, health system costs and food costs to the individual.

**Results:** 28 modelling studies on food tax policies were identified. Taxes resulted in decreased consumption of the targeted foods (e.g., -15.4% in beef/ruminant consumption, N=12 studies) and an average decrease of 8.3% in GHG emissions (N=19 studies). Using this review, we conceptualized four scenarios: a GHG weighted tax on all foods; a GHG weighted tax on food groups with the highest 50% of emissions (‘high emitters’); A GHG weighted tax on ‘high emitters’ combined with a fruit and vegetable subsidy; A 20% tax on ‘high emitters’.

The ‘GHG weighted tax on all foods’ scenario had the largest health gains and costs savings (455,800 QALYs and NZ$8.8 billion), followed by the tax-subsidy scenario (410,400 QALYs and NZ$6.4 billion). All scenarios were associated with reduced GHG emissions (between 4.2% and 7.0% of the baseline GHG emissions from food). Age standardised per capita QALYs were between 1.6 and 2.1 times higher for Māori than non-Māori.

**Conclusion:** Applying taxes that target foods with high GHG emissions has the potential to be effective for reducing GHG emissions and to result in co-benefits for population health. Combining a GHG food tax with a fruit and vegetable subsidy may help reduce the negative effects on household food expenditure of such a tax.

**Key messages:** *What is already known on this topic:* Modelling studies investigating the impact of food taxes have shown taxes aimed at high GHG emitting foods reduce consumption of ruminant meats and GHG emissions. No reviews of modelling studies of GHG motivated food taxes have been published.

*What this study adds:* Modelling studies are reviewed and summarised and used to inform modelling of four GHG motivated tax scenarios. Modelled results identify a tax/subsidy with positive impacts on population health (410,400 total or 93.2 quality adjusted life years per 1000 people over their lifetime), health system costs (NZD 6.4 billion savings), ethnic health equity (health gains were 1.6 times higher for NZ’s indigenous population, Māori than non-Māori), GHG emissions (−4.2%) and cost of diets (−0.5%).

*How this study might affect research, practice or policy:* Policymakers can use these findings in designing a food tax to benefit both climate and population health, utilising these detailed results on factors that affect population wellbeing.

## Introduction

Climate change is a major threat to human civilisation and health[1] and is increasingly recognised as a determinant of wellbeing and increasing inequities.[2] The urgent need to limit global warming by reducing greenhouse gas (GHG) emissions has gained widespread international acceptability following the United Nations Framework Convention on Climate Change and adoption of the Paris Agreement. There is a strong case for interventions targeting agricultural production as food systems are responsible for up to 29% of global anthropogenic GHG emissions.[3] One mechanism for this is through policies to change what people consume. Three quarters of global agricultural GHG emissions are associated with meat production, through land use change and enteric methane emissions[4]. Although improvements to livestock farming methods to reduce the associated GHG emissions are likely in the future,[5] there will still need to be global reductions in meat production and consumption alongside these innovations to reduce emissions.

The consumption of red and processed meat is associated with an increased risk of chronic disease. [6],[7],[8],[9] Fruits, vegetables, nuts and seeds are associated with a decreased risk of coronary heart disease (CHD), stroke, type 2 diabetes and various cancers.[10] Increased consumption of fruits, vegetables and wholegrains and reduced red and processed meat consumption could therefore reduce the detrimental effects to our long-term health and reduce the burden on the health care system.[11] Research has also demonstrated the potential climate co-benefits of these dietary changes. [12]

Taxes have been shown to reduce consumption of harmful products, such as alcohol,[13] tobacco[14] and sugar-sweetened beverages[15], and evidence shows that the more products a tax covers the more effective it is in changing people’s purchasing behaviours. It is also important to consider any potentially regressive effects of a tax and to design the tax to reduce negative outcomes, in terms of both the financial impact on households and health inequities. While GHG food taxes are internationally regarded as potential instruments to help achieve emissions reductions,[16] clear gaps in the evidence-base exist.[17, 18] The appropriateness and efficacy of taxes are likely to vary across different contexts, and the different ways food taxes can be designed and implemented will also affect the impact of a tax.

This paper aimed to first review the international literature of the effects of food taxes motivated by reducing GHG emission. We then used this evidence to design a variety of taxes aimed at reducing the purchasing, and therefore consumption, of high GHG-emitting foods. These taxes were then modelled in New Zealand, a high-income country with a western diet to be used as a case study to estimate the taxes impact on population health gain (in quality-adjusted life-years: QALYs), health system cost-savings, health inequities between Māori (NZ indigenous population) and non-Māori, GHG emissions and food costs. At present there is no NZ-specific evidence on the extent to which GHG emission taxes on food could impact health,[19-21] health inequities and health system costs.

## Methods

### Literature review

We took a systemised approach to reviewing studies on tax policies motivated by reducing GHG emissions, the query string used within each database combined search terms relating to four themes: *climate change; food systems; tax policies;* and *health*. We searched Scopus and Web of Science (WoS), limited to the past 10 years (2010-2020, search run on 8^th^ December 2020) and those written in English. Four independent searches (In Scopus and Web of Science with and without the ‘health theme’) were carried out. (See the supplementary material for additional details including box S1 for the search strategy). An iterative process was used to refine the search strategy to produce the final search terms (Box S1).

Both modelling studies and real-world evaluations were included if they:

a. illustrated a tax motivated by reducing GHG emissions
b. allocated a quantifiable tax amount to a defined food group or groups.

Studies were excluded if they:

a. included only a tax on foods which was not motivated by reducing GHG emissions
b. did not allocate a quantifiable tax amount to a defined food group or groups
c. only assessed environmental outcomes other than GHG emissions, e.g., biodiversity loss
d. only related to subsidy policies
e. only included a tax that was not specific to food groups (e.g., fuel tax or electricity tax)

All references of included studies after initial title/abstract screening were screened for inclusion. Studies within reviews identified were also screened against the exclusion and inclusion criteria. We extracted the following data from included studies: location, study design, tax justification/purpose, tax description and quantity, targeted food group/type, and outcomes including changes in price, amount of food produced and/or consumed, GHG emissions, impacts on health disparities, health system costs, and population health.

### Modelling methods

#### Dietary intake data

Dietary intake data were sourced from the most recent representative New Zealand Adult National Nutrition Survey (NZANS) and used as an estimate for baseline intake. This was conducted in 2008/09 (data acquired from the University of Otago’s Life in New Zealand Research Group who conducted the survey, through personal communication, Blakey, Smith and Parnell, 2014). Dietary data were from a single 24-hour dietary recall and are in grams (g) per food group for each of 338 food groups. Average intakes per food group were calculated for sex by ethnic groups (Māori and non-Māori).

We used an existing NZ-specific price elasticity matrix disaggregated into a 338 by 338 food group matrix to align with consumption data. We constrained total food expenditure using a total food expenditure elasticity (TFEe) of 0.75. These methods are well-established and consistent with earlier work modelling the impact of food taxes in NZ. Further detail is presented in the supplementary material and elsewhere.[22-24]

#### Modelling

The tax scenarios were applied to the baseline diet (the business-as-usual (BAU) comparator) in the dietary intervention model. The taxes influenced food purchases through price elasticities, which subsequently affected consumption. Differences in food consumption between BAU and the tax scenarios were simulated for the entire New Zealand population, alive in 2011 (N=4.4 million), using an Excel based dietary proportional multi-state life-table model (PMSLT). Outputs from this modeling were changes in daily GHG emissions per person in kgCO_2_-eq, percentage change in daily cost of diet per person, the price index of the diet (a measure of relative price changes of the total diet) and the following outputs over the life course of those alive in 2011: incremental population QALYs gained; ethnic health inequities (ratio of age adjusted per capita health gains between Māori and non-Māori) and costs or cost-savings to the health system in New Zealand dollars (NZ$). Detailed modelling methods are included in the supplementary material. See supplementary table 1 for baseline input parameters.

#### GHG emissions

The units for the carbon taxes are tonnes of kgCO_2_-eq whereby CO_2_-eq refers to a comparable unit that averages GHG emissions to equal the same global warming impact as CO_2_, using the standard GWP_100_ method (GWP_100_ is the accounting metric adopted by the Intergovernmental Panel on Climate Change in inventory guidelines), this allows for comparison between products with different levels of GHGs.

A New Zealand-specific life-cycle assessment (LCA) database was previously developed by modifying cradle to point-of-sale reference emissions estimates from an established UK database to the New Zealand context. This UK database, presented in Hoolohan et al. (2013),[25] provided per-kg cradle to point-of-sale emissions estimates for 66 food categories. It included the relative contributions of the following lifecycle stages: farming and processing; transportation; transit packaging; consumer packaging; warehouse and distribution; refrigeration; and supermarket overheads. Each NZANS food group used in modelling was matched to a NZ specific LCA (if available) or a food category from the reference LCA database and emissions estimates were assigned accordingly.

UK emissions estimates were modified to the NZ context, with efforts focused on lifecycle stages that contributed most to overall emissions and those where the NZ context was expected to differ most from the UK database (transportation and electricity usage). Further details on these methods are outlined in Drew et al 2020.[26]

CO_2_-eq per 100g of food group was used to calculate the amount of tax applied to each food group. To calculate the threshold to be used to define high GHG emitting foods (henceforth referred to as the ‘high emitters’) we averaged the CO_2_-eq per 100g of food for the 338 food groups in the NZANS: 0.46 kgCO_2_-eq/100g.

#### GHG tax scenarios

Among the various GHG food tax approaches proposed within the literature, four were selected for further investigation and corresponding scenarios were designed on the basis that they provided a range of options to help understand the impact of differently designed taxes.

The taxes designed for modelling were designed primarily to reduce GHG emissions, while exploring their potential for achieving health co-benefits and reducing health inequities between Māori and non-Māori. To explore the potentially regressive impacts of food taxes we designed one that attempts to minimise the overall financial impact of taxes on consumers by providing a compensatory subsidy for fruit and vegetables. The scenarios were informed by the results of the literature review and details of selected taxes are presented in the results section.

##### Sensitivity and further scenario analyses

Māori are disadvantaged in the main analysis as they have higher background morbidity and mortality, this results in a lesser ‘envelope’ for potential health gains. In order to value health gain in Māori the same as for non-Māori, an equity analysis was modelled in which background morbidity and mortality rates for Māori were set to non-Māori values.[27] All scenarios were rerun with no discounting so health gain in the future is valued the same as health gain in the present. A lower and upper sensitivity analysis was carried out for all scenarios modelled.

##### Patient and public involvement

No patients or public were involved in the design of this study.

## Results

### Review Results

A total of 3,564 records were identified across the four searches conducted (see figure 1). Following review, there were 28 included studies, of which 27 were modelling studies, with one cost-benefit modelling analysis.[28] The locations of these studies were the UK (5),[17, 29-32] international (4),[33-36] Spain (4),[37-40] the EU (3),[41-43] France (3),[44-46] Sweden (2),[47, 48] The Netherlands,[28] Switzerland,[49] Denmark,[18] Canada,[50] Australia,[51] Norway[52] and Belgium.[4] Seven studies also included a subsidy on food groups.[18, 28, 34, 37, 40, 46, 50] Seven had a fixed percentage tax on food products.[28, 31, 32, 38, 40, 45, 52] The remaining 21 studies had calculated tax rates based on a carbon price per unit of food.[2, 4, 17, 35, 49, 53] The carbon taxes ranged 25,000-fold from 0.01 GBP/tCO_2_-eq (0.02 NZD)[31] to 290 EUR/tCO_2_-eq (489.92 NZD).[41] The food groups targeted ranged from all food groups to just two groups.[34, 36, 40] There were 14 studies which only taxed animal products,[28, 31-34, 36, 38, 41, 42, 44-46, 48, 50] two of which also included a scenario that taxed all products.[31, 46] Supplementary table 2 summarises the key characteristics and results of the included studies.

**Figure 1:**
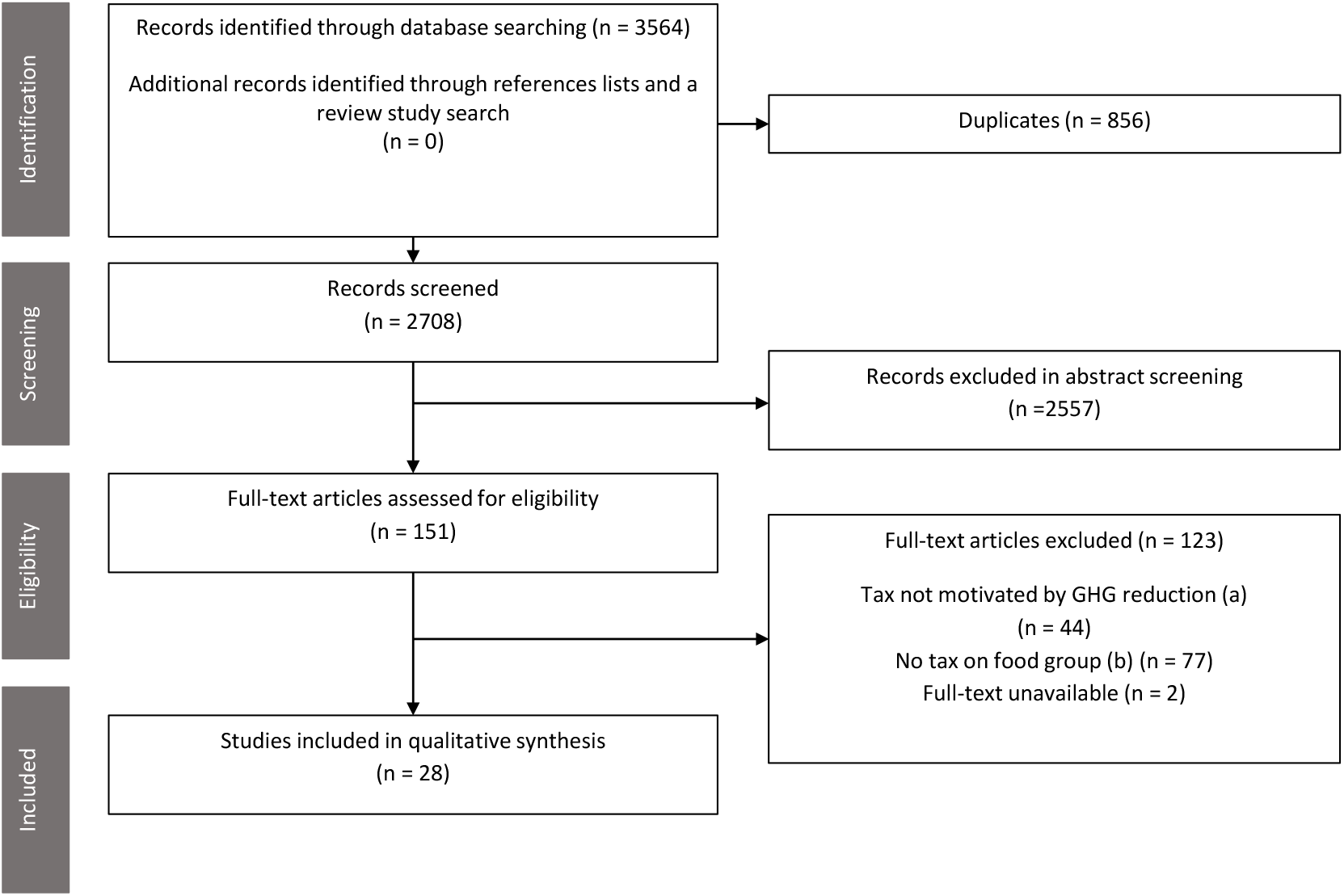
A PRISMA flow diagram following the process of study identification and eligibility screening.

Prices increased mainly for animal products, except in Dogbe et al.[37] where fish/seafood prices decreased by 5% and 15% when tax revenues generated from the taxed foods were used to subsidize lower emission foods under a 56 EUR/tCO_2_-eq (94.54 NZD) and 200 EUR/tCO_2_-eq (337.65 NZD) tax respectively. In the 21 publications reporting percentage change in the price of beef, the average price change was 28.1% (range -8.3%[40] to 90.1%[34]). The average increase in price for studies that targeted dairy products was 22.5% (N=14, range 1.9%[11] to 113.7%[41]).

Generally, modelling studies showed that taxing foods based on their GHG emissions decreases beef/ruminant consumption, with an average decrease of 15.4% (N=12 studies reporting % change in consumption) with changes ranging from an increase in consumption of 10.4%[40] to a decrease of 49.0%.[52] Due to cross-price elasticities (that represent changes in purchases of food groups that are not taxed, e.g. if they are substitutes for the taxed foods), some modelling studies showed an increase in consumption of non-alcoholic drinks and fresh fruit; pork and poultry; and snacks and other foods.[37, 42, 51] Of 9 studies that examined dietary energy intake, 8 saw a decrease,[17, 18, 30, 35, 39, 44, 46, 51] the largest being a 14.9% reduction.[46]

GHG emissions generally decreased when a tax was modelled. One study designed their tax to meet specific GHG emission reduction targets.[52] All other studies showed modelled reductions in GHG emissions ranging from 0.4%[43] under a 50 USD/tCO_2_-eq (69.46 NZD) tax to 19.4%[18] under a 760 DKK/tCO_2_-eq (174.80 NZD) tax. The average of the 19 papers reporting percentage change in GHG emissions for the relevant jurisdiction was -8.3% (when multiple scenarios were reported, the largest change was included in this calculation). In contrast, Broeks et al.[28], who modelled a 10% subsidy on fruit and vegetables in the Netherlands, found an increase of 4.5% in environmental impact based on GHG emissions, acidification, water eutrophication and land use.

Vandenberghe et al.[4] observed a saving of up to 79,800 disability-adjusted life years (DALY) for a tax of 60 EUR/tCO_2_-eq (101.88 NZD) in the modelled year in Belgium. A higher tax on all foods (96 Swiss Franc per tCO_2_-eq (151.02 NZD)) estimated a reduction of 706 DALYs per year for the Swiss population.[49] Springmann et al.[51] estimated a reduction of 49,500 DALYs or 1,620 averted deaths at a tax rate of 23 AUD/tCO_2_-eq (24.66 NZD) in Australia. 7770 deaths were modelled to be averted in a UK modelling study (tax of 2.72 GBP/tCO_2-_eq (5.17 NZD) but when this tax was modelled in combination with a subsidy for low GHG emission foods an extra 2685 deaths occurred.[17] However, lower estimates of deaths delayed or averted was seen with a similar modelled tax (2.86/tCO_2_-eq (5.44 NZD)) alone (300), combined with a subsidy on low emission foods (90), combined with a 20% SSB tax (1200) or combined with both the subsidy and SSB tax (2000).[29] Springmann et al.[35] estimated 107,000 avoided deaths at a global scale, with a tax rate of 52 USD/tCO_2_-eq (72.24 NZD).

Only two of the included studies commented on the health system impacts of a modelled GHG emission food tax. Vandenberghe et al.[4] suggested that a 30, 45 and 60 EUR/tCO_2_-eq (50.72, 76.11, and 101.88 NZD) tax in Belgium could save €256–€481 million in the modelled year in terms of medical expenditure. Broeks et al.[28] reported that meat taxes of 15% or 30%, or a 10% fruit and vegetable subsidy could save up to 7.4, 12.3, and 3.3 billion euros over 30 years, respectively.

### Modelling Results

#### Modelled GHG food tax scenarios

##### S1: GHG weighted tax, all foods

Firstly, we have chosen a conceptually simple approach where all foods are taxed based on their GHG emissions. This approach has been taken in previous modelling studies[30, 37, 49] with the levels of taxation varying between 5.40 NZD per tCO_2_-eq[30] and 337.65 NZD per tCO_2_-eq.[37]

The approach taken for this paper was to set the tax level so the ANS dietary food group ‘beef, muscle meat’ (chosen as it is 100% meat rather than a composite food group such as casserole) increased in price by 20%. This is a relatively arbitrary tax level often chosen in tax modelling and advocacy, as such we carried out sensitivity analyses which corresponded to 10% and 40% for each scenario. The amount of tax necessary to increase the price of ‘beef, muscle meat’ by 20% was $163.59/tCO_2_-eq/100g of food. This amount was applied to all food groups in the ANS and is applied per 100g of food to line up with the structure of the model. Values used in the S1(lower) and S1(upper) scenarios were $82.03/tCO_2_-eq/100g of food and $327.18/tCO_2_-eq/100g of food.

##### S2: GHG weighted tax, ‘high emitters’

The second scenario is the same as the first but targets ‘high emitters’ (emissions above 0.46 kgCO_2_-eq/100g). As in S1, the magnitude of the tax is weighted by the GHG emissions of each food group ($163.59/tCO_2_-eq/100g). This modelling approach, targeting high emitting foods only, was taken in Bonnet et al[44] and in one of the scenarios presented in Dogbe et al[37]. Values used in the S2(lower) and S2(upper) scenarios were the same as in S1.

##### S3: GHG weighted tax and subsidy

Thirdly, we combined the weighted tax outlined for S2 on ‘high emitters’, with a 20% subsidy on all fruit and vegetables. S3 aims to be approximately price neutral to reduce the negative impact on household finances of regressive taxes. Previous modelling studies have used subsidies in combination with taxes to partially offset their financial impacts on individuals[17, 18, 46] with value-added tax being removed on all foods[18] or subsidies being applied to low emitting foods[17] and fruits, vegetables and starchy foods[46]. Values used in the S3(lower) and S3(upper) scenarios were the same as in S1 for the tax and 10% for S3 (lower) and 40% for S3(upper) for the subsidy.

##### S4: Percentage tax on ‘high emitters’

As the first three scenarios may be administratively difficult to implement, we modelled a more straightforward proxy. We applied a set percentage tax on the ‘high emitters’. This approach is similar to several other modelling studies, which taxed meat (beef, pork and chicken[50], Beef and sheep meat[34, 36] or animal products[31, 38, 41]. Taxes have been set to a range of percentages or set amounts per tCO_2_-eq for specific high GHG emitting foods[34, 36, 41, 50] and one modelling study presented both approaches[31]. A 20% tax was chosen following the same rationale as S1-S3. This was the same as in Caillevat et al[45] and was one of the taxes used in Revoredo-Giha et al[31]. Values used in the S4(lower) and S4(upper) scenarios were 10% and 40% taxes. See supplementary table 3 for tax rates and target food groups for all scenarios.

#### Impacts on modelling outcomes

##### S1

Average changes in the dietary risk factors which impact on disease incidence were as follows: BMI decreased by 0.5kg/m^2^; red and processed meat intake by 10g/p/d with small decreases in vegetable and sodium intakes. Small increases were seen in intakes of fruit, sugar sweetened beverages, polyunsaturated fat and nuts (see Table 1). The largest changes in food group intake, compared to baseline, was a decrease of ‘milk’ (−22g) and ‘grains and pasta – rice only’ (−20g) (Supplementary Table 4).

**Table 1.** Population weighted change in BMI and dietary risk factors used for modelling of taxes.

| | $\Delta$ BMI | $\Delta$ Fruit<br>(g/day) | $\Delta$ Vege-<br>tables<br>(g/day) | $\Delta$ Red<br>meat<br>(g/day) | $\Delta$ Process-<br>ed meat<br>(g/day) | ΔSSB<br>(g/day) | $\Delta$ Nuts<br>and<br>seeds<br>(g/day) | $\Delta$ Sodium<br>(mg/<br>day) | $\Delta$ PUFA<br>(%<br>TE) |
| --- | --- | --- | --- | --- | --- | --- | --- | --- | --- |
| S1: GHG weighted tax, all foods | -0.48 | 3.2 | -1.6 | -4.1 | -5.5 | 1.0 | 0.2 | -52.2 | 0.1% |
| S2: GHG weighted tax, high emitters | -0.10 | 3.8 | 4.0 | -5.5 | -5.9 | 2.8 | 0.2 | -25.4 | 0.0% |
| S3: GHG weighted tax and subsidy | -0.19 | 28.4 | 50.4 | -7.4 | -7.3 | -0.1 | -0.1 | -50.8 | 0.0% |
| S4: Percentage tax on high emitters | 0.03 | 9.7 | 10.2 | -4.7 | -7.8 | 7.0 | 0.5 | -34.6 | 0.0% |

BMI: body mass index; SSB: sugar sweetened beverage; TE: total energy intake

These changes in intake led to the largest health gains (432,000 QALYs gained over the remaining lifespan of the NZ population alive in 2011, or 98 QALYs gained per 1000 people alive in 2011) and cost savings (NZ$8.2 billion) of all scenarios (Table 2). Age standardised per capita health gain for Māori was 1.8 times that of non-Māori. This ratio increased to 2.3 when the equity analysis was modelled. Health gain was higher for men than for women (ratio of 1.3). S1 also generated the greatest GHG emission reductions compared to the baseline NZ diet, approximately 0.35kgCO_2_-eq per person per day or an 7.0% reduction in average diet-generated GHG emissions (Table 3). Cost of diet increased by 3.8% with a change in the price index of 5.0%.

**Table 2.** Lifetime health impacts (in QALYs) and health system costs for GHG food taxes, for the NZ population alive in 2011 (lifetime horizon) with 3% discount rate.

|  | Non-Māori | Māori | Māori | Ethnic groups combined |  |
| --- | --- | --- | --- | --- | --- |
| | Health gains:<br>QALYs | Health gains:<br>QALYs | Equity<br>analysis[27]<br>health gains:<br>QALYs | Health gains:<br>QALYs | Net health<br>system cost<br>savings (NZ\$<br>billion) |
| **S1: GHG weighted tax, all foods** |  |  |  |  |  |
| Total | 327,300 (226,500<br>to 467,800) | 104,700 (70,700<br>to 153,400) | 138,100 (94,100<br>to 202,200) | 432,000 (298,200<br>to 615,000) | \$8.2 (5.4 to<br>12.4) |
| Men | 184,000 | 58,200 | 77,100 | 242,200 | \$4.7 |
| Women | 143,300 | 46,400 | 61,000 | 189,700 | \$3.6 |
| Per capita* | 87.7 (113.6) | 155.2 (201.3) | 204.9 (266.4) | 98.1 | \$1,866.8 |
| **S2: GHG weighted tax, highest emitters** |  |  |  |  |  |
| Total | 143,700 (92,000<br>to 219,500) | 53,200 (32,400<br>to 84,000) | 69,700 (44,200 to<br>107,600) | 196,900 (125,000<br>to 303,000) | \$3.6 (2.1 to<br>5.8) |
| Men | 84,700 | 28,800 | 37,900 | 113,500 | \$2.1 |
| Women | 59,000 | 24,400 | 31,800 | 83,300 | \$1.5 |
| Per capita* | 38.5 (49.6) | 78.9 (102.4) | 103.4 (134.4) | 44.7 | \$816.6 |
| **S3: GHG weighted tax and subsidy** |  |  |  |  |  |
| Total | 322,000 (259,800<br>to 391,300) | 88,400 (74,000<br>to 104,900) | 118,900 (99,400<br>to 141,400) | 410,400 (336,200<br>to 492,000) | \$6.4 (4.9 to<br>8.2) |
| Men | 175,500 | 44,500 | 59,800 | 220,000 | \$3.6 |
| Women | 146,500 | 44,000 | 59,100 | 190,400 | \$2.8 |
| Per capita* | 86.3 (106.5) | 131.2 (170.6) | 176.3 (229.6) | 93.2 | \$1,451.8 |
| **S4: Percentage tax on highest emitters** |  |  |  |  |  |
| Total | 118,500 (4,900 to<br>296,200) | 34,300 (-8,300<br>to 100,100) | 46,600 (-8,000 to<br>126,900) | 152,800 (-3,000<br>to 396,600) | \$2.4 (-.7 to<br>7.2) |
| Men | 63,200 | 13,200 | 18,500 | 76,400 | \$1.2 |
| Women | 55,200 | 21,100 | 28,200 | 76,400 | \$1.2 |
| Per capita* | 31.7 (39.9) | 50.9 (66.7) | 69.2 (90.7) | 34.7 | \$549.8 |

\*per capita results: QALYs/1000 people & \$/person

**Table 3.** Health, health system cost and GHG emission impacts of sensitivity and further scenario analyses around the base case for each scenario.

| Sensitivity/scenario analyses | Health gains: QALYs (millions) | Net health system cost savings (NZ\$ billion) | Change in kgCO<sub>2</sub>-eq per person per day | Cost of diet per person per day (% change from baseline diets) |
| --- | --- | --- | --- | --- |
| **S1: GHG weighted tax, all foods** |  |  |  |  |
| Base case analysis* | 0.44 | \$8.5 | -0.35 | 3.8% |
| S1 (lower) | 0.24 | \$4.6 | -0.18 | 1.9% |
| S1 (upper) | 0.75 | \$14.2 | -0.67 | 7.6% |
| Undiscounted | 1.62 | \$23.7 | | |
| **S2: GHG weighted tax, high emitters** |  |  |  |  |
| Base case analysis* | 0.20 | \$3.7 | -0.21 | 1.9% |
| S2 (lower) | 0.11 | \$2.1 | -0.11 | 1.0% |
| S2 (upper) | 0.33 | \$5.9 | -0.41 | 3.9% |
| Undiscounted | 0.74 | \$10.0 | | |
| **S3: GHG weighted tax and subsidy** |  |  |  |  |
| Base case analysis* | 0.42 | \$6.5 | -0.21 | -0.5% |
| S3 (lower) | 0.25 | \$4.1 | -0.12 | -0.3% |
| S3 (upper) | 0.52 | \$6.7 | -0.30 | -1.0% |
| Undiscounted | 1.55 | \$16.8 | | |
| **S4: Percentage tax on high emitters** |  |  |  |  |
| Base case analysis* | 0.16 | \$2.5 | -0.22 | 4.7% |
| S4 (lower) | 0.11 | \$1.9 | -0.12 | 2.4% |
| S4 (upper) | 0.07 | \$0.0 | -0.37 | 9.5% |
| Undiscounted | 0.59 | \$6.5 | | |

\*3% discounting, no discounting applied to GHG emissions

##### S2

There was a small decrease in BMI (−0.1kg/m^2^), red meat and processed meat (−11g/p/d), sodium, (−25mg/p/d) and small increases in fruit and vegetables (8g/p/d) and SSBs (3g/p/d). The largest decreases in food group intake, compared to baseline, was a decrease in ‘beef & veal’ (−5g), ‘sausage & processed meats’ (−4g) and ‘bread-based dishes’ (−4g).

Health gains and cost savings were approximately half that of S1 (196,900 QALYs and NZ$3.6 billion). Age standardised per capita health gain for Māori was 2.1 times, and 2.7 times when the equity analysis was applied, that of non-Māori. Health gain was higher for men than for women (ratio of 1.4). S2 saved approximately 0.21kgCO_2_-eq of GHG emissions per person per day (4.3% of the baseline diet). Cost of diet increased by 2.5% with a change in the price index of 1.9%.

##### S3

BMI decreased by an average of 0.2kg/m^2^, red and processed meat by 15g/p/d and sodium by 51mg/p/d. Fruit and vegetable intake increased by 28g and 50g respectively. The largest changes in food group intake, compared to baseline, was a decrease of ‘non-alcoholic beverages’ (−24g) alongside the increases in fruit and vegetable intake.

An increase of 410,400 QALYs was seen with associated cost-savings to the health system of $6.4 billion. Age standardised per capita health gain for Māori was 1.6 times that of non-Māori and 2.2 higher when the equity analysis was modelled. Health gain was higher for men than for women (ratio of 1.2). The difference in consumption between the baseline diet and S3 saved an average of 0.21kgCO_2-_eq of GHG emissions per person per day (4.2% of baseline diets). Cost of diet decreased by 0.5% with a change in the price index of -0.8%.

##### S4

There was an increase in fruit and vegetable intake (20g) and SSB intake (7g) and a small average increase in BMI of 0.03kg/m^2^. Sodium decreased by 35mg/p/d, and red and processed meat by 13g/p/d. The largest changes in food group intake, compared to baseline, was a decrease of ‘bread-based dishes’ (−13g) and ‘fish and seafood’ (−5g).

QALY gains were 152,800 and health system cost savings were $2.4 billion. Age standardised per capita health gain for Māori was 1.7 times that of non-Māori and 2.3 times higher when the equity analysis was modelled. Health gain was 1.6 times higher for Māori women than Māori men but 1.1 times higher for non-Māori men than in non-Māori women. S4 saved approximately 0.22kgCO_2-_eq of GHG emissions per person per day (4.5% of baseline diets). Cost of diet increased by 4.7% with a change in the price index of 6.2%.

##### Sensitivity and further scenario analyses

Table 3 presents the sensitivity and further scenario analysis results. Health gain was approximately 3.7 times greater when results were not discounted over time for all scenarios. Health gain in the lower scenarios (equivalent to a 10% increase in the price of ‘beef, muscle meat’) was between 0.5 and 0.7 of the base case health gain for all scenarios. Health gain in the upper scenarios (equivalent to a 40% increase in the price of ‘beef, muscle meat’) was between 0.4 of the health gains (due to an increase in BMI in S4 (upper)) and 1.7 times greater than the base case scenarios. Health system cost savings were approximately 2.7 times greater when undiscounted but otherwise mirrored these patterns. GHG savings in the lower scenarios were approximately half of the base case scenarios. GHG savings in the upper scenarios savings were between 1.4 (S3) and 2.0 (S2) times greater than the base case scenarios.

Figure 2 plots health gain against GHG emission reductions to visually represent health and climate co-benefits and includes the main 4 scenarios with their upper and lower sensitivity analyses. S1(upper) gives the most health gain and GHG emission reductions of all taxes by a clear margin. This is followed by S3 scenarios and then S2 scenarios. S4 is the only set of scenarios which does not show a linear trend between the lower, main and upper scenarios with the health gain for S4 (upper) being lower than for S4 (lower). This is due to the change in BMI being -0.03, 0.03 and 0.37 for S4 (lower), S4 and S4 (upper) respectively.

**Figure 2.**
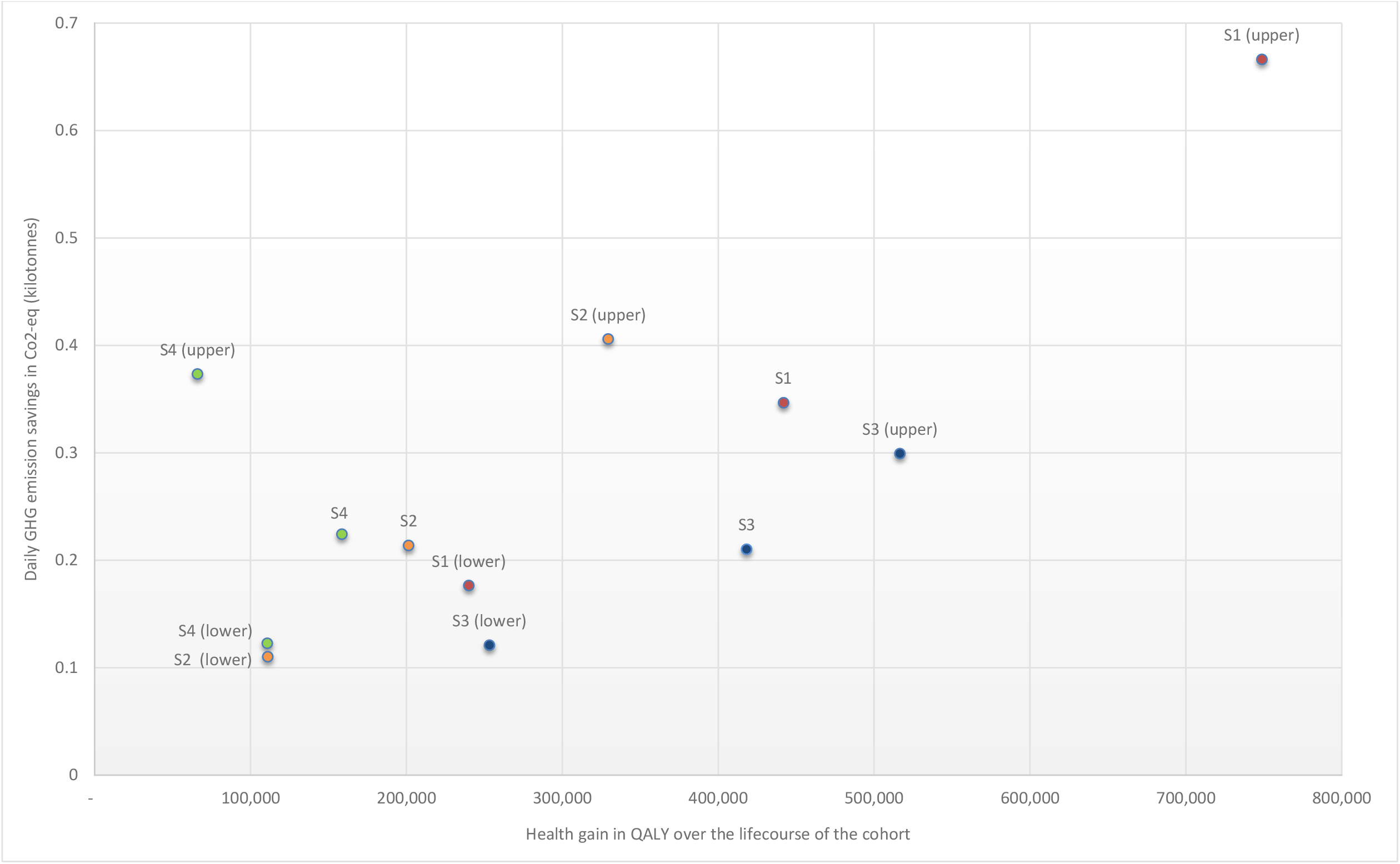
GHG emission impact against health impact of the four main scenarios and their upper (equivalent to a 40% increase in the price of ‘beef, muscle meat’) and lower (equivalent to a 10% increase in the price of ‘beef, muscle meat’) sensitivity analyses.

Supplementary table 5 shows QALYs and costs/cost savings that occur in the first 10 and 20 years of the taxes. Between 4% and 5% of lifetime QALYs accrue in the first ten years and between 18% and 19% in the first twenty years of the taxes. Cost savings were between 9% and 12% of lifetime cost savings and between 30% and 34% in the first ten and twenty years of the taxes.

## Discussion

### Main findings

The review found 28 modelling studies of food tax policies motivated by reducing GHG emissions. Most of these studies calculated tax rates based on a carbon price per unit of food using a wide range of tax rates. Studies consistently showed that the increased price of targeted foods decreased their consumption and decreased total GHG emissions.

We developed a range of tax scenarios based on the review. The largest health gains and cost savings occurred in the scenario where a tax was applied to all foods, weighted by GHG emissions closely followed by the tax subsidy scenario. The changes in dietary risk factors responsible for these health gains and cost savings were increases in fruits and vegetables (up to almost 80g for the tax subsidy scenario) and small decreases in red and processed meat and sodium. Most of the health gain, however, was due to the decrease in average BMI across the population from the decrease in the energy content of the taxed diets. All scenarios were associated with GHG savings (between 4.2% and 7.0% of baseline diets).

Indigenous Māori in NZ are disproportionately affected by the health effects of diet, particularly foods and nutrients linked to cardiovascular disease and type 2 diabetes.[54] This is a result of both higher consumption and wider structural injustices. NZ has an obligation under te Tiriti o Waitangi (the constitutional Treaty between the British Crown and Māori) to support Māori health and policies that affect diets need to consider how they can reduce health inequities between Māori and non-Māori. Health gain was between 1.6 and 2.1 times higher for Māori than non-Māori for these scenarios (using age standardised per capita QALYs) so these taxes have the potential to reduce ethnic health inequities.

It is also important to consider impacts on household costs when considering tax policy design. Food costs to the individual increased by between 1.9% and 4.7% for the tax only scenarios and decreased by 0.5% for the tax subsidy scenario. The GHG tax on all food scenario has the largest health gain, cost savings, GHG impact and second highest potential impact on health equity. However, combining a GHG tax with a fruit and vegetable subsidy may be the best approach to employ considering the large health gain and cost savings, reductions in GHG emissions and food costs, improvements in fruit and vegetable consumption and health equity and practical implementation considerations.

### Strengths and limitations

The dietary data for the modelled baseline consumption are taken from the latest Adult National Nutrition survey, carried out in 2008/09, and consumption in New Zealand may have changed over the last 12 years. Data from the NZ health survey show a steady decrease in adult fruit and vegetable consumption over this time period (https://minhealthnz.shinyapps.io/nz-health-survey-2020-21-annual-data-explorer/). The modelled impacts on health may therefore be conservative. The base year of the PMSLT model used was 2011 with trends on disease incidence, case fatality and remission out until 2026. Disease rates may have changed since 2011, potentially altering the impact of these taxes.

There is uncertainty around the extent to which price elasticities can predict future changes in purchasing which, in this modelling, is applied to consumption data to estimate the change in BMI. The price elasticities matrix that we used was generated using Bayesian priors from previously published New Zealand food price elasticities.[24] The applied TFEe method aimed to limit implausible changes in food intake that may be generated through breaches in econometric assumptions of PE estimation.

This study uses CO_2_ equivalents as the metric for GHG emissions. There is an argument to be made that methane should be reported separately to carbon dioxide and nitrous oxide due to their different length of warming potentials, and the specific methane targets developed at the global level (e.g., “Global Methane Pledge” at the COP26 U.N. Climate Summit, 2021). Three of the modelled taxes have targeted food groups with high GHG emissions, those with more than the average CO_2_-eq per 100g of food for the 338 food groups in the NZANS. This does not take into consideration the amount of these foods consumed in NZ and it may be more efficient to focus taxes on commonly consumed high emitters. This was not explored in this paper.

### Policy implications

The results of the review clearly show positive impacts on consumption and GHG emissions of modelled GHG food taxes, though these would not be enough alone to meet obligations for food emissions reductions. The modelling shows health and climate co-benefits from four different GHG food pricing policies. There would be practical challenges translating these taxes to a real-world setting, including likely opposition from food producers and industry, how GHGs would be measured, the metrics used to equivalise methane, and how product and producer specific the tax should be.

It also needs to be acknowledged that there are particular limitations in NZ to taxing domestic consumption where a large proportion of high-emitting foods are produced for export (e.g. 89% of bovine meat and 87% of mutton and goat meat in 2019[55]). Much greater reductions in GHG emissions would likely be achieved by targeting agricultural production in NZ. Equity implications of taxing individual consumers rather than pricing emissions at the point of production are also important to consider. However, considering the scope and urgency of the reductions in GHG emissions needed, multiple policies that have synergistic effects on food production and consumption will need to be implemented, ideally including policies with health co-benefits such as a consumption tax on high GHG emitting foods. Additionally, food producers remain sensitive to their domestic consumers, and taxing food consumption may increase social pressure to address GHG emissions from total agricultural production.

### Conclusions

Applying consumption taxes that target foods with high GHG emissions associated with their production has the potential to result in population health and climate co-benefits alongside potential savings to the health system and reductions in ethnic health inequities. Whilst NZ is used as a case study, findings are likely to be of relevance to other high-income countries. Policymakers need to weigh up the benefits of such a consumer food tax against other options that are likely to result in improved diets and reduced food system GHG emissions. A consumption tax combined with a subsidy on fruit and vegetables could be considered to minimise the impact on household food costs.

## Supporting information

Cheers checklist

## Data Availability

All data produced in the present study are available upon reasonable request to the authors

## Acknowledgements

National nutrition survey data (2008/09) acquired from the University of Otago’s Life in New Zealand Research Group who conducted the survey, through personal communication (Blakey, Smith and Parnell, 2014). Access to the data used in this study was provided by Statistics New Zealand under conditions designed to keep individual information secure in accordance with requirements of the Statistics Act 1975. The opinions presented are those of the author(s) and do not necessarily represent an official view of Statistics New Zealand.

## Contributor statement

CC led the conceptualization and design of the study, led the modelling and wrote the first draft of the paper. IM, AlM, AnM, TB and CnM contributed to the conceptualization and design of the study. CC, TB, NN and AnM contributed to building the models. IM carried out the literature review, supervised by CC and AlM. JD developed the GHG database. All authors contributed to the interpretation of results and the drafting and revision of the manuscript.

## References

1. Costello A, Abbas M, Allen A, Ball S, Bell S, Bellamy R, Friel S, Groce N, Johnson A, Kett M: Managing the health effects of climate change: lancet and University College London Institute for Global Health Commission. Lancet 2009, 373(9676):1693–1733.

2. Whitmee S, Haines A, Beyrer C, Boltz F, Capon AG, de Souza Dias BF, Ezeh A, Frumkin H, Gong P, Head P: Safeguarding human health in the Anthropocene epoch: report of The Rockefeller Foundation–Lancet Commission on planetary health. Lancet 2015, 386(10007):1973–2028.

3. Vermeulen SJ, Campbell BM, Ingram JS: Climate change and food systems. Annu Rev Environ Resour 2012, 37.

4. Vandenberghe D, Albrecht J: Tackling the chronic disease burden: are there co-benefits from climate policy measures? Eur J Health Econ 2018, 19(9):1259–1283.

5. Panchasara H, Samrat NH, Islam N: Greenhouse gas emissions trends and mitigation measures in australian agriculture sector—a review. Agriculture 2021, 11(2):85.

6. Abete I, Romaguera D, Vieira AR, Lopez de Munain A, Norat T: Association between total, processed, red and white meat consumption and all-cause, CVD and IHD mortality: a meta-analysis of cohort studies. Br J Nutr 2014, 112(05):762–775.

7. Feskens EJ, Sluik D, van Woudenbergh GJ: Meat consumption, diabetes, and its complications. Curr diab rep 2013, 13(2):298–306.

8. International Agency for Research on Cancer (World Health Organisation): IARC Monographs evaluate consumption of red meat and processed meat. In., No. 240 edn. Lyon, France; 2015: 2.

9. Lim SS, Vos T, Flaxman AD, Danaei G, Shibuya K, Adair-Rohani H, AlMazroa MA, Amann M, Anderson HR, Andrews KG: A comparative risk assessment of burden of disease and injury attributable to 67 risk factors and risk factor clusters in 21 regions, 1990–2010: a systematic analysis for the Global Burden of Disease Study 2010. Lancet 2013, 380(9859):2224–2260.

10. Forouzanfar MH, Alexander L, Anderson HR, Bachman VF, Biryukov S, Brauer M, Burnett R, Casey D, Coates MM, Cohen A: Global, regional, and national comparative risk assessment of 79 behavioural, environmental and occupational, and metabolic risks or clusters of risks in 188 countries, 1990–2013: a systematic analysis for the Global Burden of Disease Study 2013. Lancet 2015, 386(10010):2287–2323.

11. Springmann M, Mason-D’Croz D, Robinson S, Wiebe K, Godfray HCJ, Rayner M, Scarborough P: Health-motivated taxes on red and processed meat: A modelling study on optimal tax levels and associated health impacts. PloS One 2018, 13(11).

12. Willett W, Rockström J, Loken B, Springmann M, Lang T, Vermeulen S, Garnett T, Tilman D, DeClerck F, Wood A: Food in the Anthropocene: the EAT–Lancet Commission on healthy diets from sustainable food systems. Lancet 2019, 393(10170):447–492.

13. Elder RW, Lawrence B, Ferguson A, Naimi TS, Brewer RD, Chattopadhyay SK, Toomey TL, Fielding JE, Services TFoCP: The effectiveness of tax policy interventions for reducing excessive alcohol consumption and related harms. Am J Prev Med 2010, 38(2):217–229.

14. Bank W: Curbing the epidemic: governments and the economics of tobacco control. Tob Control 1999, 8(2):196–201.

15. Teng AM, Jones AC, Mizdrak A, Signal L, Genç M, Wilson N: Impact of sugar-sweetened beverage taxes on purchases and dietary intake: Systematic review and meta-analysis. Obes Rev 2019, 20(9):1187–1204.

16. Golub AA, Henderson BB, Hertel TW, Gerber PJ, Rose SK, Sohngen B: Global climate policy impacts on livestock, land use, livelihoods, and food security. Proc Natl Acad Sci 2013, 110(52):20894–20899.

17. Briggs AD, Kehlbacher A, Tiffin R, Garnett T, Rayner M, Scarborough P: Assessing the impact on chronic disease of incorporating the societal cost of greenhouse gases into the price of food: an econometric and comparative risk assessment modelling study. BMJ Open 2013, 3(10):e003543.

18. Edjabou LD, Smed S: The effect of using consumption taxes on foods to promote climate friendly diets–The case of Denmark. Food Policy 2013, 39:84–96.

19. Macdiarmid JI, Kyle J, Horgan GW, Loe J, Fyfe C, Johnstone A, McNeill G: Sustainable diets for the future: can we contribute to reducing greenhouse gas emissions by eating a healthy diet? Am J Clin Nutr 2012, 96(3):632–639.

20. Reynolds CJ, Buckley JD, Weinstein P, Boland J: Are the dietary guidelines for meat, fat, fruit and vegetable consumption appropriate for environmental sustainability? A review of the literature. Nutrients 2014, 6(6):2251–2265.

21. Wilson N, Nghiem N, Ni Mhurchu C, Eyles H, Baker MG, Blakely T: Foods and dietary patterns that are healthy, low-cost, and environmentally sustainable: a case study of optimization modeling for New Zealand. PLoS One 2013, 8(3):e59648.

22. Blakely T, Nghiem N, Genc M, Mizdrak A, Cobiac L, Mhurchu CN, Swinburn B, Scarborough P, Cleghorn C: Modelling the health impact of food taxes and subsidies with price elasticities: The case for additional scaling of food consumption using the total food expenditure elasticity. PloS One 2020, 15(3):e0230506.

23. Cleghorn C, Blakely T, Nghiem N, Mizdrak A, Wilson N: Technical report for BODE³ intervention and DIET MSLT models, Version 1. Burden of Disease Epidemiology, Equity and Cost-Effectiveness Programme. Technical Report no. 16. In. Edited by Department of Public Health UoO, Wellington. Wellington; 2017.

24. Jacobi L, Nghiem N, Ramírez-Hassan As, Blakely T: Food Price Elasticities for Policy Interventions: Estimates from a Virtual Supermarket Experiment in a Multistage Demand Analysis with (Expert) Prior Information. Econ Rec 2021.

25. Hoolohan C, Berners-Lee M, McKinstry-West J, Hewitt C: Mitigating the greenhouse gas emissions embodied in food through realistic consumer choices. Energy Policy 2013, 63:1065–1074.

26. Drew J, Cleghorn C, Macmillan A, Mizdrak A: Healthy and Climate-Friendly Eating Patterns in the New Zealand Context. Environ Health Perspect 2020, 128(1):017007.

27. McLeod M, Blakely T, Kvizhinadze G, Harris R: Why equal treatment is not always equitable: the impact of existing ethnic health inequalities in cost-effectiveness modeling. Popul Health metr 2014, 12(1):15.

28. Broeks MJ, Biesbroek S, Over EA, van Gils PF, Toxopeus I, Beukers MH, Temme EH: A social cost-benefit analysis of meat taxation and a fruit and vegetables subsidy for a healthy and sustainable food consumption in the Netherlands. BMC Public Health 2020, 20:1–12.

29. Briggs AD, Kehlbacher A, Tiffin R, Scarborough P: Simulating the impact on health of internalising the cost of carbon in food prices combined with a tax on sugar-sweetened beverages. BMC Public Health 2015, 16(1):1–14.

30. Kehlbacher A, Tiffin R, Briggs A, Berners-Lee M, Scarborough P: The distributional and nutritional impacts and mitigation potential of emission-based food taxes in the UK. Clim Change 2016, 137(1):121–141.

31. Revoredo-Giha C, Chalmers N, Akaichi F: Simulating the impact of carbon taxes on greenhouse gas emission and nutrition in the UK. Sustainability 2018, 10(1):134.

32. Chalmers NG, Revoredo-Giha C, Shackley S: Socioeconomic effects of reducing household carbon footprints through meat consumption taxes. J Food Prod Mark 2016, 22(2):258–277.

33. Key N, Tallard G: Mitigating methane emissions from livestock: a global analysis of sectoral policies. Clim Change 2012, 112(2):387–414.

34. Revell BJ: One man’s meat… 2050? Ruminations on future meat demand in the context of global warming. J Agric Econ 2015, 66(3):573–614.

35. Springmann M, Mason-D’Croz D, Robinson S, Wiebe K, Godfray HCJ, Rayner M, Scarborough P: Mitigation potential and global health impacts from emissions pricing of food commodities. Nat Clim Chang 2016.

36. Revell B: Meat and Milk Consumption 2050: the Potential for Demand-side Solutions to Greenhouse Gas Emissions Reduction. EuroChoices 2015, 14(3):4–11.

37. Dogbe W, Gil JM: Effectiveness of a carbon tax to promote a climate-friendly food consumption. Food Policy 2018.

38. Forero-Cantor G, Ribal J, Sanjuán N: Levying carbon footprint taxes on animal-sourced foods. A case study in Spain. J Clean Prod 2020, 243:118668.

39. García-Muros X, Markandya A, Romero-Jordán D, González-Eguino M: The distributional effects of carbon-based food taxes. J Clean Prod 2017, 140:996–1006.

40. Markandya A, Galarraga I, Abadie LM, Lucas J, Spadaro JV: What Role Can Taxes and Subsidies Play in Changing Diets? Finanz-Archiv: Zeitschrift für das Gesamte Finanzwesen 2016, 72(2):175.

41. Jansson T, Säll S: Environmental consumption taxes on animal food products to mitigate Greenhouse gas emissions from the European Union. Clim Chang Econ 2018, 9(04):1850009.

42. Wirsenius S, Hedenus F, Mohlin K: Greenhouse gas taxes on animal food products: rationale, tax scheme and climate mitigation effects. Clim Change 2011, 108(1):159–184.

43. Zech KM, Schneider UA: Carbon leakage and limited efficiency of greenhouse gas taxes on food products. J Clean Prod 2019, 213:99–103.

44. Bonnet C, Bouamra-Mechemache Z, Corre T: An environmental tax towards more sustainable food: empirical evidence of the consumption of animal products in France. Ecol Econ 2018, 147:48–61.

45. Caillavet F, Fadhuile A, Nichèle V: Taxing animal-based foods for sustainability: environmental, nutritional and social perspectives in France. Eur Rev Agric Econ 2016, 43(4):537–560.

46. Caillavet F, Fadhuile A, Nichèle V: Assessing the distributional effects of carbon taxes on food: Inequalities and nutritional insights in France. Ecol Econ 2019, 163:20–31.

47. Moberg E, Andersson MW, Säll S, Hansson P-A, Röös E: Determining the climate impact of food for use in a climate tax—design of a consistent and transparent model. Int J Life Cycle Assess 2019, 24(9):1715–1728.

48. Säll S, Gren M: Effects of an environmental tax on meat and dairy consumption in Sweden. Food Policy 2015, 55:41–53.

49. Chen C, Chaudhary A, Mathys A: Dietary change scenarios and implications for environmental, nutrition, human health and economic dimensions of food sustainability. Nutrients 2019, 11(4):856.

50. Slade P: The effects of pricing Canadian livestock emissions. Can J Agric Econ/Revue canadienne d’agroeconomie 2018, 66(2):305–329.

51. Springmann M, Sacks G, Ananthapavan J, Scarborough P: Carbon pricing of food in Australia: an analysis of the health, environmental and public finance impacts. Aust N Z J Public Health 2018, 42(6):523–529.

52. Abadie L, Galarraga I, Milford A, Gustavsen G: Using food taxes and subsidies to achieve emission reduction targets in Norway. J Clean Prod 2016, 134:280–297.

53. Wilcock RJ, Monaghan RM, Quinn JM, Campbell AM, Thorrold BS, Duncan MJ, McGowan AW, Betteridge K: Land-use impacts and water quality targets in the intensive dairying catchment of the Toenepi Stream, New Zealand. N Z J Mar Freshwater Res 2006, 40(1):123–140.

54. Lucero AA, Lambrick DM, Faulkner JA, Fryer S, Tarrant MA, Poudevigne M, Williams MA, Stoner L: Modifiable cardiovascular disease risk factors among indigenous populations. Adv Prev Med 2014, 2014.

55. Food Balance Sheets. [http://www.fao.org/faostat/en/#data/FBS]

