## Supplementary material for "Can a greenhouse gas emissions tax on food also be healthy and equitable? A systematized review and modelling study from Aotearoa New Zealand"

#### Additional review methods

##### Supplementary Box 1: Search Strategy

Databases: Scopus and WoS (2010 -2020) each with two searches (with and without the health concept).

Search terms: Article titles, abstract and keywords

All four concepts (food systems, tax policy, climate change, and health)

- At least one of: food*: meat*: diet*: agricultur*
  AND
- At least one of: tax: fiscal: "pric* polic*"
  AND
- At least one of: "climate change": greenhouse: sustain*: environment*
  AND
- At least one of: health: disease*: disparit*: equit*

Four databases were explored (OVID, Scopus, Google Scholar, and Web of Science (WoS) before identifying Scopus and WoS as the most useful with the largest reach. They identified the most validation articles (out of a pool of 40) which had been previously identified as relevant. Furthermore, these databases include literature from medicine and social sciences which are in line with the subjects of this project: climate sciences, economics, nutrition, and public health. Ryyan, an online systematic review website, was used to combine the search results, remove exact duplicates, and analyse in the title and abstract screening stage. A pre-determined inclusion and exclusion criteria was developed.

Data was extracted for the review (*Description*: Location; population; design; justification for tax rate; tax description; tax quantity; food group tax applied to. *Results:* changes seen in demand/production; changes seen in GHG emissions; food substitutions; equity impacts; health system cost impacts; population health impacts) and to inform the modelling presented in this paper (Jurisdiction; tax description; tax amount; food groups targeted by tax; tax rate converted to NZD and the tax amount applied to each Adult Nutrition Survey (ANS) food group in absolute or percentage terms).

### Additional modelling methods

#### Price elasticities

A price elasticity matrix (23 food groups by 23) was generated using a linear almost-ideal demand system. This included Bayesian priors for demand equation coefficients which were generated from a previously published New Zealand food price elasticities matrix[1, 2]. More detail on the price elasticity methods is presented elsewhere.[3, 4] There are important differences between products within these broad 23 food groups in GHG emissions (e.g. between wheat flour and rice within the 'grains and pasta' food group), and it was necessary to allow for shifts in purchasing within the broad food groups. We therefore disaggregated foods and their price elasticities into a 338 by 338 food group matrix[5], to align with consumption data in the NZANS.

Applying price elasticities matrices from one study population to another, where food consumption patterns and prices differ, might breach the econometric assumptions inherent in PE matrix estimation. This could result in underestimation or overestimation of intervention total food purchasing and therefore implausible changes in modelled energy intake and BMI. We therefore constrained total food expenditure using a total food expenditure elasticity (TFEe) of 0.75.[6]

We used two New Zealand studies[7, 8] and international studies to set a beta distribution for the TFEe to use in Monte Carlo simulations that returned a mean of 0.75. Using a TFEe of 0.75 means that for each 1% increase in total food prices, consumers increase their expenditure on food by 0.75%. There is a partial compensation, presumably at the expense of some other aspect of the household budget (e.g., reduced spending on transport or recreation). We evenly rescale all food purchasing to achieve a 0.75% increase in expenditure for each 1% increase in the total price of food. More detail on these methods is presented elsewhere.[6]

#### Modelling

Differences between BAU and the tax scenarios were simulated for the entire New Zealand population, alive in 2011 (N=4.4 million), using an Excel based dietary proportional multi-state life-table model (PMSLT). The NZ population broken down by sex, age and ethnicity (Māori, Indigenous population, and non-Māori) was modeled out to death or until year 2121 in the PMSLT. The structure and 'business as usual' (BAU) inputs for this model are described in detail in the model's technical report[5]. Costs and savings to the health system were modeled using a lifetime horizon. The taxes were modeled as if implemented in the base year (2011) and kept in place indefinitely.

The BAU model uses projected all-cause mortality and morbidity rates by sex and age, and separately for Māori and non-Māori ethnic groups. Running alongside this main life-table were 18 diet or body mass index (BMI) related disease life-tables, where proportions of the population simultaneously resided: CHD, stroke, type 2 diabetes, osteoarthritis, dental caries, and multiple cancers (i.e., endometrial, head and neck, kidney, liver, lung, esophageal, pancreatic, stomach, thyroid, colorectal, breast, ovarian and gallbladder). These contain incidence, prevalence and case-fatality and remission (the latter in cancers only) in 2011. The proportion of the New Zealand population in each disease life-table was a function of the disease incidence, case-fatality and remission (cancers only), except for dental caries, where only incidence was modelled. Future trends in cancer incidence, case-fatality and remission were specified using regression estimates of trends from historic data. Trends in other diseases were obtained from the New Zealand Burden of Disease Study (NZBDS)[9].

Morbidity was quantified, by sex, age and ethnic groups, for each disease using the years of life lived with disability (YLDs) from the NZ Burden of Disease Study (BDS), divided by the population count to give prevalent YLDs. Disability weights from the Global Burden of Disease Study 2010 were used to estimate the health status valuation of these YLDs [10].

The intervention effect was captured through changes in dietary intake and BMI (resulting from changes in energy intake[11]) due to the food taxes. The change in dietary risk factors and BMI was then combined with relative risks for the associations between risk factor and diseases through population impact fractions (percentage reductions in future diet/BMI-related disease incidence) that alter the disease incidence in the relevant disease life-tables. All disease input parameters were specified by sex, age and ethnicity unless stated differently (see Supplementary Table 1).

Time lags from change in dietary risk factor intake/BMI to change in disease incidence were allowed for by using the average change in risk factors over a previous window of time of: zero to five years for cardiovascular disease, diabetes and osteoarthritis, and 10 to 30 years for cancers. Probabilistic distributions about the boundaries (five, 10 and 30 years) were also specified[5].

Health system costs (sex and age-specific) were calculated in 2011 NZ$ using individually-linked data for publicly funded (and some privately funded) health events occurring in 2006-10, including hospitalizations, inpatient procedures, outpatients, pharmaceuticals, laboratories and expected primary care usage. Costs were sourced from the New Zealand Health Tracker database for all diseases except diabetes (Virtual Diabetes Register[12]) and dental caries (a weighted estimate of published treatment costs). The cohort was assigned an (sex and age-specific) annual health system cost of a citizen without a diet/BMI-related disease and not in the last six months of their life.[13] Additional disease-specific excess costs were assigned to people for those in the first year of diagnosis, last six months of life if dying of the given disease, and otherwise prevalent (or incident in the case of dental caries) cases of each disease in the model. Costs were modeled over the lifetime of the cohort, including costs both related and unrelated to the diet/BMI-related diseases modeled (i.e. increased longevity due to the taxes resulted in increased health system costs for some cohort members). Intervention costs were included in the modelling i.e. the cost of a law (NZ$3.5 million)[14] to introduce new legislation on food taxes.

Microsoft Excel using an Ersatz add-in was used to run 2000 simulations of each of the tax scenarios with uncertainty. Each of these simulations involved a random draw from the probability density function about those parameters specified with uncertainty (Supplementary table 1). The model used 3% discounting which reduced the reported outputs by 3% each subsequent year, valuing health gains and costs/cost savings in the short term more than gains in the long term. The modelling takes a health system perspective.

**Supplementary table 1: Baseline input parameter**

| Key parameter | Source/ Application to Model | Expected Value and uncertainty | Distribution/ Heterogeneity |
|---|---|---|---|
| Baseline population count | Stats New Zealand (SNZ) population estimates for 2011. | Nil uncertainty. | Sex<br>Age<br>Ethnicity |
| All-cause mortality rates | SNZ mortality rates for 2011. | Nil uncertainty. | Sex<br>Age<br>Ethnicity |
| Disease-specific incidence, prevalence, case-fatality rates, and remission rates | For each disease, a coherent set of incidence rates, prevalence, case-fatality rates (CFR), and remission rates (zero for non-cancers) were estimated using DISMOD II using data from the Ministry of Health, NZ burden of disease study (NZBDS) and HealthTracker. | Uncertainty: rates all +/- 5% standard deviation (SD). | Log-normal<br>Sex<br>Age<br>Ethnicity |
| Disease trends | Trends are applied to incidence, case-fatality, and remission rates until 2026 and then kept constant for the remainder of the lifetimes of the modeled population. | Uncertainty +/- 0.5% absolute change.<br>Diabetes: Uncertainty +/- 1.5% absolute change. | Normal<br><br>Sex<br>Ethnicity |
| Total morbidity per capita in 2011 | The per capita rate of years of life lived with disability (YLD) from the NZBDS. | Uncertainty +/- 10% SD. | Log-normal<br>Sex<br>Age<br>Ethnicity |
| Disease morbidity rate per capita | Each disease was assigned an age and sex specific disability rate (DR) equal to YLDs for that disease (scaled down to adjust for comorbidities) from the 2006 NZBDS projected forward to 2011. This was divided by the disease prevalence. This was then assigned to the proportion of the cohort in each disease state. | Uncertainty: +/- 10% SD. | Normal<br><br>Sex<br>Age |
| Health system costs | Linked health data (hospitalizations, inpatient procedures, outpatients, pharmaceuticals, laboratories, and expected primary care usage) for all New Zealanders had unit costs assigned to each event (for the period 2006–2010), and then health system costs (NZD2011) were estimated. | Estimated at SD= ±10% of the point estimate. | Gamma<br>Sex<br>Age |
| Time-lags for intervention effect | It takes time for a change in dietary risk factors to impact on disease incidence. As there are no precise data on just how long these are we have used wide windows of time-lags with wide uncertainty. For cancers the time-lag is assumed to range between 10 and 30 years. For coronary heart disease (CHD), stroke, diabetes and osteoarthritis (the | Uncertainty: +/- 20% SD. | Normal |

| Key parameter | Source/ Application to Model | Expected Value and uncertainty | Distribution/ Heterogeneity |
|---|---|---|---|
| | non-cancers), the time-lag is assumed to be between 0 and 5 years. | | |
| TMREL | The Theoretical Minimum Risk Exposure Level (TMREL) is the level of risk exposure that is theoretically possible and minimizes overall risk and is derived from the latest Global Burden of Disease 2013 study. [15] It allows us to estimate how much of the disease burden could be lowered by shifting the distribution of a risk factor to the level that would lead to the greatest improvement in population health. | Uncertainty: Uniform distribution between 0 and 1 | Uniform |
| Height of the NZ adult population (for BMI calculations) | Mean and SD of height from NZ Adult Nutrition Survey 2008/09[16] | Uncertainty using reported SD. | Normal<br>Sex<br>Ethnicity |
| Intervention costs | The cost of a law (NZ$3.5 million)[14] to introduce new legislation on food taxes. | 95% UI: NZ$2.0 to NZ$6.2 million | Gamma |

### Additional review results

**Supplementary Table 2: A descriptive summary table of 28 studies on the tax effectiveness on reducing GHG emissions and improving health.**

| ***Study/jurisdiction*** | ***Tax amount*** | ***Food groups*** | ***Findings*** |
|---|---|---|---|
| Abadie 2016[17] (Norway) | Four emissions reductions targets (2.5%, 5%, 7.5%, and 10%). Tax amounts were calculated for each of the 16 food groups for each target. | All food and drinks (16 groups) | The largest decreases in food quantities consumed to meet the 2.5% to 10% emission reduction targets were for ruminants (-14 to -49%) and cheese (-4% to -28%). The largest increases were for other foods (4% to 67%), fish (8% to 42%) and poultry (6% to 39%). |
| Bonnet 2018[18] (France) | A) 56 EUR/t$CO_2$-eq (94.54 NZD/t$CO_2$-eq).<br>B) 200 EUR/t$CO_2$-eq (337.65 NZD/tCO2-eq). | 1) all animal products, 2) beef, veal, lamb, and sheep products, 3) beef only. | Consumption of animal products reduced between A3) -0.3g and B1) -23.3g per person per day.<br><br>GHG emissions ($CO_2$-eq per year) reduced between A3) -1.1% and B1) -6.1%. |
| Briggs 2013[19] (UK) | A) 2.72 GBP/t$CO_2$e/100g (5.17 NZD/t$CO_2$-eq/100g) for high GHG emission foods.<br>B) A + subsidy for low GHG emission foods. | All food and drinks (29 groups) | Largest decrease in consumption with beef (A) -14.2% & B) -13.7%) and lamb (A) -12.1% and B) -13.9%).<br><br>GHG emissions (kt$CO_2$-eq) reduced: A) -18 700 kt$CO_2$-eq, B) -15 200.<br><br>A) 7770 deaths averted. B) 2685 extra deaths. |
| Briggs 2015[20] (UK) | A) 2.86/t$CO_2$-eq (5.44 NZD/t$CO_2$-eq) to food groups emissions > 0.36 kg$CO_2$-eq.<br>B) same but subsidising groups with emissions < 0.36 kg$CO_2$-eq/100g.<br>C) A with 20% SSB tax. D) B with 20% SSB tax. | All food and drinks (32 groups). | Greatest reductions in purchased amounts were for beef (-20.5% to -21.3%) and 'not low calorie' soft-drinks (C) -10.5% and D) -26.2%).<br><br>GHG emissions (kt$CO_2$-eq) reduced: A) -18 900, B) -17 100, C) -18 500, D) -16 500.<br><br>A) 300, B) 90, C) 1200, D) 2000 deaths delayed or averted. |
| Broeks 2020[21] (NL) | A) 15% meat tax. B) 30% meat tax C) 10% fruit and vegetable subsidy | Meat (red meat, processed meat, and poultry) and fruit and vegetables. | Reduction in daily meat consumption (A) -8.8g and B) -16.7g). Increase in daily fruit and vegetable consumption (C) 11g).<br><br>GHG emission (t$CO_2$-eq) changes for 2048: A) -900,000 C) 90,000.<br><br>Net societal benefit: A) €3100 to 7400 million. B) €4000 to 12,300. C) €1800 to 3300 million over 30 years (when a QALY value of €50,000 was applied). |

| | | | |
|---|---|---|---|
| Caillevat 2016[22] (France) | A) 20% tax on high GHG emission products. B) 20% tax on five food groups with highest GHG emissions and fat content. | A) Animal products (8 groups) B) Animal products (5 groups) | GHG emissions (household g$CO_2$-eq) decreased: A) -7.5%, B) -7.0%. |
| Caillevat 2019[23] (France) | A) 56 euros per t$CO_2$-eq (94.54 NZD) and B) 140 (235.44 NZD). Applied in each scenario (1-3). | 1) Taxes all food groups 2) Taxes animal products 3) Taxes animal products and subsidises fruit, vegetables and starchy foods. | Largest increase in price was for animal-based foods, high in fat (1A) 9.5%, 1B) 23.3%). Largest decrease in price was for Fresh fruits and vegetables (3A) -4.9%, 3B) -14.9%).<br><br>GHG emissions (household g$CO_2$-eq) decreased: 1A) -6.2%, 1B) -15.5%, 2A) -2.2%, 2B) -5.5%, 3A) -1.1%, 3B) -1.8%). |
| Chalmers 2016[24] (UK, Scotland) | The tax rate based on the carbon footprint and the marginal damage cost of the meat categories: Beef 13.0%, chicken 3.2%, pork 6.3%, turkey 4.2% and sheep 12.0%. | Beef, chicken, pork, turkey and sheet | Meat related GHG emissions ($CO_2$-eq) reduced by 10.5%. |
| Chen 2019[25] (Switzerland) | 96 Swiss Franc per t$CO_2$-eq (151.02 NZD) | All foods and milk (28 groups) | Largest change in consumption was a reduction of dairy products by 12 g/person/day. All other changes were less than 5g.<br><br>GHG emissions (kg$CO_2$eq/person/day) decreased by 2.2.<br><br>Reduced DALY: 706 per year for the Swiss population. |
| Dogbe 2018[26] (Spain) | Uncompensated scenarios (taxes were proportional to the food groups carbon footprint):<br>A) 56 EUR/t$CO_2$-eq (94.54 NZD)<br>B) 200 EUR/tCO2-eq (337.65 NZD)<br>Compensated revenue neutral scenarios (tax revenues generated from the taxed foods were used to subsidize lower emission foods):<br>C) 56 EUR/t$CO_2$-eq (94.54 NZD)<br>D) 200 EUR/t$CO_2$-eq (337.65 NZD) | A) and B) All foods and milk (16 food groups)<br>C) and D): 7 food groups with higher GHG-emissions: all meats, milk and dairy products, cheese and composite dishes (the other 9 food groups were subsidised). | Largest price increases were for composite dishes (C)5%, A) 15%, B & D) 55%) and beef, veal and lamb (A & C)12% and B & D) 44%). Largest price decreases in the compensated scenarios were for starchy roots, tubers, legumes, nuts and oilseeds (C) -8%, D) -27%) and sugar and confectionary and prepared desserts (C) -7%, D) -25%). All animal-based foods increased in price apart from the fish and seafood category which decreased in the compensated scenarios (C) -5%, D) -19%).<br><br>Pork consumption decreased the most and the residual and snacks and other foods consumption increased the most in scenario D.<br><br>GHG emissions decreased by 2.0%-6.4% in the compensated scenarios. |
| Edjabou 2013[27] (Denmark) | 1A) all foods taxed at 0.26 DKK /kg $CO_2$-eq (0.059 NZD) 1B) all foods taxed at 0.76 DKK /kg $CO_2$-eq (0.17 NZD). 2A&B: Same, but tax | All foods and milk (23 groups) | Beef showed the largest price increase (1A) 11.1%, 1B) 32.4%, 2A) 8.5%, 2B) 25.3%). Other dairy showed the smallest price increase in the first 2 scenarios (1A) 0.4%, 1B) 1.0%) and one of the largest decreases in price in the revenue neutral scenarios (2A) -2.3%, 2B) -6.1%). |

| | | | |
|---|---|---|---|
| | revenue from that carbon tax is matched to a VAT reduction. | | GHG emissions (kg $CO_2$-eq/person/year) decreased: 1A) -7.9%, 1B) -19.4%, 2A) -3.4%, 2B) -8.8%. |
| Forero-Cantor 2020[28] (Spain) | 10%, 12.5%, 15%, 17.5% & 20% tax on each food group, applied separately. | Beef, chicken, eggs, fish, pork, lamb, turkey | Taxes may be highly effective in reducing GHG emissions when applied on fish (e.g., a 15% tax on fish resulted in a total net reduction of 17.82 million tons of $CO_2$-eq), moderately effective on beef, eggs and lamb, ineffective on chicken and turkey, and counterproductive on pork meat. |
| Garcia-Muros 2017[29] (Spain) | A) 25 EUR/tCO2-eq (42.33 NZD). B) 50 EUR/tCO2-eq (84.66 NZD). C) Scenario B but exempting cereals, fruits, milk and vegetables. | A & B) All food groups (13 groups) C) Beef, pork, poultry, fish, dairy, eggs, potatoes, oil, sugar. | GHG emissions decreased: A) -3.8% and B) -7.6% (total reductions not reported for C). |
| Jansson and Säll 2018[30] (EU) | A) 16 EUR/t$CO_2$-eq (27.03NZD), B) 60 EUR/t$CO_2$-eq (101.36NZD), C) 290 EUR/t$CO_2$-eq (489.92 NZD) | Animal products (15 groups) | The largest percent increases in consumer price were for butter (A) 5.1% B) 20.3% C) 113.7%) and then whole milk powder (A) 4.4% B) 16.8% C) 83.3%).<br><br>Agricultural emission decreased for EU: A) -0.5% B) -1.47% C) -4.93% and globally: A) -0.06% B) -0.20% C) -0.75%. This equates to reductions (Mt$C0_2$-eq) of: EU: A) -2.0 B) -5.9 C) -19.9 and globally: A) -3.3 B) -10.4 C) -38.5. |
| Kehlbacher 2016[31] (UK) | 2.841 GBP/t$CO_2$-eq (5.40 NZD) | A) All food and drinks (29 groups)<br>B) High GHG emission foods (9 groups) | Beef exhibited the largest price increase (10.6% to 12.4%), followed by milk (Scenario A only, 8.7% to 11.9%).<br><br>Change in consumption varied widely by socio-economic class. The largest decrease in consumption is seen in 'intermediate, small employers & own account workers' for sweets (-22.1%) and sugar etc (-28.0%).<br><br>GHG emissions ($CO_2$-eq) decreased: A) -6.3% and B) -4.3%. |
| Key 2012[32] (International) | 30 USD/tCO2-eq. (182.43 NZD) for methane emissions only (varies across countries and commodities depending on average methane emissions per unit). | Meat and milk (6 groups) | Change in production varied from -4.7% for beef to 1.1% for poultry for Annex 1 countries and from -6.5% for beef to 0.3% for poultry for non-Annex 1 countries.<br><br>Total methane emissions (for 2013) decreased by -3.9% and -4.5% in Annex 1 countries and non-Annex 1 countries respectively. |
| Markandya 2016[33] (Spain) | Tax rate set to reach a A) 25% improvement and B) 20% improvement in reaching nutritional targets from the baseline diet. | All foods and drinks (39 groups) | Largest decrease in consumption is for nuts and seeds (A) -37.3% and B) -32.7%) followed by margarine and low-fat spread (A) -36.7% and B) -34.4%). Both these food groups were taxed at the largest rate of 30% and 25%. Largest increase in consumption is for processed potatoes (A) 61.9% |

| | | | |
|---|---|---|---|
| | A) maximum of 30% and B) maximum of 25% tax or subsidy. | | and B) 48.9%) followed by biscuits (A) 54.6% and B) 41.0%). Both these food groups were subsidised at the largest rate of 30% and 25%.<br><br>GHG emissions ($CO_2$/person/day) decreased by 13% for food taxes to meet nutritional targets. |
| Moberg 2019[34] (Sweden) | Tax rate applied based on the different greenhouse gases: 1120 SEK/t$CO_2$ (NZD 186.89), 35,797 SEK/t$CH_4$ (NZD 5966.79) and 438,762 SEK/t$N_2O$ (NZD 73134.61). | All foods sold in Sweden. | n/a: tax design only. |
| Revell 2015[35] (International) | A) USD 80/t$CO_2$-eq (111.34 NZD) per tonne of ruminant meat consumption emissions in developed economies from 2010 to 2050. B) The same tax applied universally. | Ruminant meats | Compared to projected global ruminant meat consumption in 2050 beef and ovine meat consumption (combined) is estimated to decrease by A) -1 Mt and B) -6 Mt.<br><br>GHG emissions (Mt$CO_2$-eq) decreased each decade in comparison to no tax: 369 (2020), 345 (2030), 329 (2040), 249 (2050). |
| Revell 2015[36] (International) | A) USD 80/t$CO_2$-eq (111.34 NZD) in developed economy regions (North America, Europe and Oceania). B) The same tax applied universally. | Beef and sheep meat | A) Global meat consumption reduced by 2 Mt (-0.4%). B) Beef and sheep/goat meat consumption reduced by just under 7 Mt (-5.4%).<br><br>A) Global GHG emissions decreased by: A) <-1%, B) -3%, both compared with projected baseline emissions in 2050. |
| Revoredo-Giha 2018[37] (UK) | A) Ad-valorem tax (set to 20% for food groupings 1 to 3 and ranged between 5% and 30% under grouping 4, depending on the kg$CO_2$-eq associated with the food groups). B) Ad-valorem tax based on the kg$CO_2$-eq associated with the food groups with 3 different prices for carbon: current average Emission Trading System (ETS) price: 0.0128 GBP/kg$CO_2$-eq (0.024 NZD), mean social cost of carbon: 0.0427 GBP/kg$CO_2$-eq (0.081 NZD) and (long term EU projection of carbon price: 0.1709 GBP/kg$CO_2$-eq (0.33 NZD)<br><br>C) As A) but "compensated" where the total revenue received from the tax was | 1) Beef and veal, other meats, not preserved 2) All meats and eggs 3) All animal-based products 4) All products | Largest decreases in GHG emissions within each scenario A) to D) was A4) -9.3%, B4) (0.1709 GBP/kg$CO_2$-eq) -18.7%, C3) -4.9%, D3) (0.1709 GBP/kg$CO_2$-eq) -15.7%.<br><br>The change in consumption associated with these scenarios with the largest GHG emission savings were: A4) cheese (-22.3%), pork (-18.8%) and grains and grain based products (-17.7%) B4) (0.1709 GBP/kg$CO_2$-eq) beef, veal and lamb (-39.4%), milk, dairy products and dairy product imitates (-39.2%) and grain and grain based products (-34.0%) C3) cheese (-23.9%), milk, dairy products and dairy product imitates (-20.7%) and poultry, eggs and other fresh meat (-18.3%) D3) (0.1709 GBP/kg$CO_2$-eq) milk, dairy products and dairy product imitates (-41.5%), beef, veal and lamb (-38.9%) and cheese (-22.1%). |

| | | | |
|---|---|---|---|
| | redistributed back amongst products. D) As B) but "compensated" | | |
| Säll and Gren 2015[38] (Sweden) | The tax level for each meat product was derived from the environmental damage caused by GHG, nitrogen, ammonia and phosphorus at the production stage of that meat and dairy product in Sweden. The tax corresponded to between 8.9% and 33.3% of the respective price per kg product in 2009. | Beef, pork, chicken, milk, fermented products, cream, cheese | Greatest price changes were for beef (33.3%), milk (22.4%) with the lowest price change for chicken (8.9%).<br><br>The largest percentage reductions in demand were for beef (-19%, -4.7kg/person), followed by pork (-8%, -2.9kg/person) and cheese (-6.3%, -1.2kg/person).<br><br>GHG emissions ($CO_2$-eq) decreased by approximately 12%. |
| Slade 2018[39] (Canada) | 50 CAD/t$CO_2$-eq (54.66 NZD). | Beef, pork, chicken | Price increase of: Beef: 8% (CAD 0.77/kg), pork: 1% (CAD 0.06/kg), chicken: 0.3% (CAD 0.02/kg).<br><br>Beef consumption is reduced by 2.6%, pork and chicken consumption are increased by 0.4% and 1.1%, respectively.<br><br>GHG emissions (t$CO_2$-eq) decreased by 342 000 (2.2% of domestic GHG emissions from livestock). |
| Springmann 2016[40] (International) | 52 USD/t$CO_2$-eq (72.24 NZD) | A) all foods, B) exclude fruit and vegetables, staples, and legumes from taxation, C) animal-based foods (meats, eggs, milk), D) red meat (beef, lamb, pork), E) beef. | Total GHG emissions (Mt$CO_2$-eq) decreased: A) -1,003, B) -962, C) -959, D) -689, E) -657.<br><br>Avoided deaths globally (2020): A) 107 000, B) 140 000, C) 137 000, D) 145 000,<br>E) 91 000. |
| Springmann 2018[41] (Australia) | 23 AUD/t$CO_2$-eq (24.66 NZD). | All food and drinks (23 groups) | Largest price increases were 17% for beef sausages, 6–7% for lamb, beef, and the category of other meat. Largest decreases in consumption were for beef sausages (-11.2%), other meat (-7.9%) and lamb (-7.5%).<br><br>Food-related GHG emissions (Mt$CO_2$-eq) decreased by 2.3 (5.8% of food-related GHG emissions).<br><br>49,500 DALYs prevented. |

| | | | |
|---|---|---|---|
| Vandenberghe 2018[42] (Belgium) | A) 30 EUR/$tCO_2$-eq (50.72 NZD), B) 45 EUR/$tCO_2$-eq (76.11 NZD) C) 60 EUR/$tCO_2$-eq (101.88 NZD). | Cereal, dessert, beef, pork, poultry, processed meat, fish, dairy and egg, fruits and vegetables. | Meat consumption reduced: A) -5.7%, B) -8.4%, C) -10.8%.<br><br>Total GHG emissions reduced: A) -3.3%, B) -4.9%, C) -6.5%.<br><br>DALY prevented: A) 42,300, B) 61,700, C) 79,800 DALY. Savings to the health system: A) €256 million, B) €373 million, C) €481 million. |
| Wirsenius 2011[43] (EU) | 60 EUR/$tCO_2$-eq (101.48 NZD) | Ruminant meat, pork, poultry, eggs and dairy products. | Price increases: Ruminant meat (16%), pig meat (5%), poultry meat (4%), milk (9%), and eggs (5%). Ruminant meat consumption was reduced by 15%. Pig and poultry (substituting ruminant meat) was increased by 1% and 7% respectively.<br><br>EU agricultural GHG emissions were decreased by 7% (30 Mt $CO_2$eq /year) excluding land use change. 80% of this is related to a decrease in ruminant meat consumption. |
| Zech and Schneider 2019[44] (EU) | 50 USD/$tCO_2$-eq (69.46 NZD) | All foods and milk (12 groups) | Highest price increase is for beef (0.63 USD/kg) and mutton/goat meat (0.61 USD/kg).<br><br>Meat consumption reduces by 2% in the EU. EU meat exports increase by 41% but a perceived demand reduction within the EU means global meat prices decrease. Demand reduction in the EU is offset by increased exports. Rest of the world's meat demand increases by 0.2% and production decreases by 0.1%.<br><br>EU GHG emission decrease by 0.4% (1670 kt/$CO_2$-eq). Increased exports out of the EU would decrease meat production in the rest of the world and yield additional GHG emission reductions equivalent to 0.68% of the EU's agricultural GHG emissions (2800 kt $CO_2$-eq). 43% of the GHG reduction indicted by a reduction in domestic consumption is lost through emission leakage. Global GHG emission reduction of 4465 kt$CO_2$-eq. |

Currencies: AUD: Australian dollars; CAD: Canadian dollars; DKK: Danish krone; EUR: Euros; GBP: Great Britain pounds; NZD: New Zealand dollars; SEK: Swedish Krona; USD: United Stages dollars.

tCO2-eq: tonnes CO2 equivalents; Mt: million tonnes; DALY: Disability adjusted life year; SSB: Sugar-sweetened beverage; GHG: Greenhouse gas.

### Additional modelling results:

**Supplementary table 3. Details of modelled tax scenarios**

| *Main Scenario* | *Tax amount* | *Tax amount lower sensitivity analysis* | *Tax amount upper sensitivity analysis* | *Food groups* |
|---|---|---|---|---|
| S1: GHG weighted, all foods | \$163.59/t$CO_2$-eq/100g of food | \$82.03/t$CO_2$-eq/100g of food | \$327.18/t$CO_2$-eq/100g of food | All 338 food groups in the ANS* |
| S2: GHG weighted tax, high emitters | \$163.59/t$CO_2$-eq/100g of food | \$82.03/t$CO_2$-eq/100g of food | \$327.18/t$CO_2$-eq/100g of food | Food groups with higher than the average** $CO_2$-eq based on $CO_2$-eq/100g (91 food groups) |
| S3: GHG weighted tax and subsidy | \$163.59/t$CO_2$-eq/100g of food<br>Subsidy: 20% | \$82.03/t$CO_2$-eq/100g of food<br>Subsidy: 10% | \$327.18/t$CO_2$-eq/100g of food<br>Subsidy: 40% | Food groups with higher than the average** $CO_2$-eq based on $CO_2$-eq/100g (91 food groups) receive the tax. Fruits and vegetables receive the subsidy. |
| S4: Percentage tax on highest emitters | 20% | 10% | 40% | Food groups with higher than the average** $CO_2$-eq based on $CO_2$-eq/100g (91 food groups) |

*Water and artificial sweeteners have been assigned 0 GHG emissions for all scenarios

**Average is calculated as the average of the GHG emissions associated with 100 grams of each of the 338 food groups in the ANS: 0.46 kgCO2e/100g of food.

**Supplementary table 4. Food group intake at baseline and for each scenario**

| Food group | Baseline (grams/ day) | Daily Gram intake in food tax scenarios (difference from baseline in grams) | | | |
|---|---|---|---|---|---|
| | | S1: GHG weighted tax, all foods | S2: GHG weighted tax, highest emitters | S3: GHG weighted tax plus fruit & vegetable subsidy | S4: Percentage tax on highest emitters |
| Sausage & processed meats | 21 | 17 (-4) | 17 (-4) | 16 (-5) | 17 (-4) |
| Eggs & egg dishes | 20 | 18 (-2) | 18 (-2) | 17 (-2) | 15 (-4) |
| Bread based dishes | 55 | 51 (-4) | 50 (-4) | 49 (-6) | 42 (-13) |
| Beef & veal | 42 | 38 (-3) | 37 (-5) | 36 (-6) | 38 (-4) |
| Other meat | 2 | 2 (0) | 2 (0) | 2 (0) | 2 (0) |
| Lamb/mutton | 9 | 9 (0) | 9 (-1) | 8 (-1) | 9 (-1) |
| Fish/seafood | 34 | 33 (-2) | 32 (-2) | 32 (-3) | 29 (-5) |
| Sugar/sweets | 26 | 25 (0) | 24 (-2) | 23 (-3) | 22 (-3) |
| Pork | 25 | 25 (0) | 24 (-1) | 23 (-2) | 22 (-3) |
| Cheese | 12 | 12 (0) | 11 (0) | 11 (-1) | 10 (-1) |
| Poultry | 46 | 45 (-1) | 44 (-2) | 43 (-3) | 44 (-2) |
| Fats & oils | 1 | 1 (0) | 1 (0) | 1 (0) | 1 (0) |
| Butter & margarine | 11 | 10 (0) | 11 (0) | 11 (0) | 11 (0) |
| Grains and Pasta - rice only | 51 | 31 (-20) | 52 (1) | 51 (0) | 53 (2) |
| Grains and Pasta - all other | 61 | 58 (-2) | 62 (1) | 60 (0) | 63 (2) |
| Dairy products | 39 | 37 (-2) | 39 (1) | 38 (0) | 40 (1) |
| Milk | 188 | 166 (-22) | 191 (3) | 186 (-1) | 195 (7) |
| Soups & stocks | 34 | 32 (-2) | 34 (1) | 34 (0) | 35 (2) |
| Nuts & seeds | 6 | 5 (0) | 6 (0) | 5 (0) | 6 (0) |
| Fruit | 151 | 154 (3) | 154 (4) | 178 (28) | 160 (9) |
| Vegetables | 153 | 151 (-2) | 156 (4) | 202 (50) | 162 (10) |
| Breakfast cereals | 26 | 23 (-2) | 26 (1) | 26 (0) | 27 (2) |
| Potatoes, kumara, & taro | 107 | 110 (3) | 110 (3) | 109 (2) | 114 (7) |
| Bread | 95 | 95 (1) | 97 (3) | 94 (0) | 101 (6) |
| Non-alcoholic beverages | 1710 | 1,746 (35) | 1,760 (50) | 1,687 (-24) | 1,833 (123) |
| Biscuits | 13 | 13 (1) | 13 (0) | 13 (0) | 14 (1) |
| Cakes & muffins | 26 | 27 (1) | 27 (1) | 26 (0) | 29 (2) |
| Puddings/desserts | 11 | 11 (0) | 11 (0) | 11 (0) | 12 (1) |
| Savoury sauces & condiments | 23 | 24 (0) | 24 (1) | 23 (0) | 25 (2) |
| Pies & pasties | 24 | 24 (0) | 25 (1) | 24 (0) | 26 (2) |
| Snack foods | 3 | 3 (0) | 3 (0) | 3 (0) | 4 (0) |
| Snack bars | 4 | 4 (0) | 4 (0) | 4 (0) | 4 (0) |

**Supplementary table 5. Health impact and cost savings to the health system in the next 10 and 20 years**

| | Impact in subsequent 10 years | | Impact in subsequent 20 years | |
|---|---|---|---|---|
| | Health gains: QALYs | Net health system cost savings (NZ$ million) | Health gains: QALYs | Net health system cost savings (NZ$ million) |
| S1: GHG weighted tax, all foods | 18,300 (11,700 to 27,400) | $747 (485 to 1127) | 78,500 (53,600 to 112,700) | $2456 (1633 to 3656) |
| S2: GHG weighted tax, highest emitters | 8,370 (5,070 to 13,500) | $349 (212 to 548) | 35,700 (22,300 to 55,800) | $1116 (680 to 1750) |
| S3: GHG weighted tax and subsidy | 18,420 (13,570 to 24,400) | $744 (546 to 979) | 76,100 (61,100 to 94,300) | $2186 (1667 to 2813) |
| S4: Percentage tax on highest emitters | 6,350 (-450 to 16,930) | $280 (4 to 710) | 27,400 (-1,600 to 70,700) | $832 (-83 to 2251) |

### References


1. Mhurchu CN, Eyles H, Schilling C, Yang Q, Kaye-Blake W, Genç M, Blakely T: Food prices and consumer demand: differences across income levels and ethnic groups. PloS One 2013, 8(10):e75934.
2. Ni Mhurchu C, Eyles H, Genc M, Scarborough P, Rayner M, Mizdrak A, Nnoaham K, Blakely T: Effects of health-related food taxes and subsidies on mortality from diet-related disease in New Zealand: an econometric-epidemiologic modelling study. PLoS One 2015, 10(7):e0128477.
3. Blakely T, Cleghorn C, Mizdrak A, Waterlander W, Nghiem N, Swinburn B, Wilson N, Mhurchu CN: The effect of food taxes and subsidies on population health and health costs: a modelling study. Lancet Public Health 2020, 5(7):e404-e413.
4. Jacobi L, Nghiem N, Ramírez-Hassan As, Blakely T: Food Price Elasticities for Policy Interventions: Estimates from a Virtual Supermarket Experiment in a Multistage Demand Analysis with (Expert) Prior Information. Econ Rec 2021.
5. Cleghorn C, Blakely T, Nghiem N, Mizdrak A, Wilson N: Technical report for BODE$^3$ intervention and DIET MSLT models, Version 1. Burden of Disease Epidemiology, Equity and Cost-Effectiveness Programme. Technical Report no. 16. In. Edited by Department of Public Health UoO, Wellington. Wellington; 2017.
6. Blakely T, Nghiem N, Genc M, Mizdrak A, Cobiac L, Mhurchu CN, Swinburn B, Scarborough P, Cleghorn C: Modelling the health impact of food taxes and subsidies with price elasticities: The case for additional scaling of food consumption using the total food expenditure elasticity. PloS One 2020, 15(3):e0230506.
7. Michelini C: New Zealand household consumption patterns 1983–1992: An application of the almost-ideal-demand-system. N Z Econ Pap 1999, 33(2):15-26.
8. Michelini C, Chatterjee S: Demographic variables in demand systems: An analysis of New Zealand household expenditure 1984–1992. N Z Econ Pap 1997, 31(2):153-173.
9. Ministry of Health: Health Loss in New Zealand: A report from the New Zealand Burden of Diseases, Injuries and Risk Factors Study, 2006–2016. In. Wellington: Ministry of Health; 2013.
10. Salomon JA, Vos T, Hogan DR, Gagnon M, Naghavi M, Mokdad A, Begum N, Shah R, Karyana M, Kosen S et al: Common values in assessing health outcomes from disease and injury: disability weights measurement study for the Global Burden of Disease Study 2010. Lancet 2012, 380(9859):2129-2143.
11. Hall KD, Sacks G, Chandramohan D, Chow CC, Wang YC, Gortmaker SL, Swinburn BA: Quantification of the effect of energy imbalance on bodyweight. Lancet 2011, 378(9793):826-837.
12. Kvizhinadze G, Nghiem N, Atkinson J, Blakely T: Cost Off-Sets Used in BODE3 Multistate Lifetable Models Burden of Disease Epidemiology, Equity and Cost-Effectiveness Programme (BODE3). Technical Report: Number 15. In. Edited by University of Otago W. Wellington; 2016.
13. van Baal PHM, Feenstra TL, Polder JJ, Hoogenveen RT, Brouwer WBF: Economic evaluation and the postponement of health care costs. Health Econ 2011, 20(4):432-445.
14. Wilson N, Nghiem N, Foster R, Cobiac L, Blakely T: Estimating the cost of new public health legislation. Bull World Health Org 2012, 90:532-539.
15. Forouzanfar MH, Alexander L, Anderson HR, Bachman VF, Biryukov S, Brauer M, Burnett R, Casey D, Coates MM, Cohen A: Global, regional, and national comparative risk assessment of 79 behavioural, environmental and occupational, and metabolic risks or clusters of risks in 188 countries, 1990–2013: a systematic analysis for the Global Burden of Disease Study 2013. Lancet 2015, 386(10010):2287-2323.

16. University of Otago, Ministry of Health: A Focus on Nutrition: Key findings of the 2008/09 New Zealand Adult Nutrition Survey. In. Wellington Ministry of Health; 2011.
17. Abadie L, Galarraga I, Milford A, Gustavsen G: Using food taxes and subsidies to achieve emission reduction targets in Norway. J Clean Prod 2016, 134:280-297.
18. Bonnet C, Bouamra-Mechemache Z, Corre T: An environmental tax towards more sustainable food: empirical evidence of the consumption of animal products in France. Ecol Econ 2018, 147:48-61.
19. Briggs AD, Kehlbacher A, Tiffin R, Garnett T, Rayner M, Scarborough P: Assessing the impact on chronic disease of incorporating the societal cost of greenhouse gases into the price of food: an econometric and comparative risk assessment modelling study. BMJ Open 2013, 3(10):e003543.
20. Briggs AD, Kehlbacher A, Tiffin R, Scarborough P: Simulating the impact on health of internalising the cost of carbon in food prices combined with a tax on sugar-sweetened beverages. BMC Public Health 2015, 16(1):1-14.
21. Broeks MJ, Biesbroek S, Over EA, van Gils PF, Toxopeus I, Beukers MH, Temme EH: A social cost-benefit analysis of meat taxation and a fruit and vegetables subsidy for a healthy and sustainable food consumption in the Netherlands. BMC Public Health 2020, 20:1-12.
22. Caillavet F, Fadhuile A, Nichèle V: Taxing animal-based foods for sustainability: environmental, nutritional and social perspectives in France. Eur Rev Agric Econ 2016, 43(4):537-560.
23. Caillavet F, Fadhuile A, Nichèle V: Assessing the distributional effects of carbon taxes on food: Inequalities and nutritional insights in France. Ecol Econ 2019, 163:20-31.
24. Chalmers NG, Revoredo-Giha C, Shackley S: Socioeconomic effects of reducing household carbon footprints through meat consumption taxes. J Food Prod Mark 2016, 22(2):258-277.
25. Chen C, Chaudhary A, Mathys A: Dietary change scenarios and implications for environmental, nutrition, human health and economic dimensions of food sustainability. Nutrients 2019, 11(4):856.
26. Dogbe W, Gil JM: Effectiveness of a carbon tax to promote a climate-friendly food consumption. Food Policy 2018.
27. Edjabou LD, Smed S: The effect of using consumption taxes on foods to promote climate friendly diets–The case of Denmark. Food Policy 2013, 39:84-96.
28. Forero-Cantor G, Ribal J, Sanjuán N: Levying carbon footprint taxes on animal-sourced foods. A case study in Spain. J Clean Prod 2020, 243:118668.
29. García-Muros X, Markandya A, Romero-Jordán D, González-Eguino M: The distributional effects of carbon-based food taxes. J Clean Prod 2017, 140:996-1006.
30. Jansson T, Säll S: Environmental consumption taxes on animal food products to mitigate Greenhouse gas emissions from the European Union. Clim Chang Econ 2018, 9(04):1850009.
31. Kehlbacher A, Tiffin R, Briggs A, Berners-Lee M, Scarborough P: The distributional and nutritional impacts and mitigation potential of emission-based food taxes in the UK. Clim Change 2016, 137(1):121-141.
32. Key N, Tallard G: Mitigating methane emissions from livestock: a global analysis of sectoral policies. Clim Change 2012, 112(2):387-414.
33. Markandya A, Galarraga I, Abadie LM, Lucas J, Spadaro JV: What Role Can Taxes and Subsidies Play in Changing Diets? Finanz-Archiv: Zeitschrift für das Gesamte Finanzwesen 2016, 72(2):175.
34. Moberg E, Andersson MW, Säll S, Hansson P-A, Röös E: Determining the climate impact of food for use in a climate tax—design of a consistent and transparent model. Int J Life Cycle Assess 2019, 24(9):1715-1728.
35. Revell BJ: One man's meat… 2050? Ruminations on future meat demand in the context of global warming. J Agric Econ 2015, 66(3):573-614.

36. Revell B: Meat and Milk Consumption 2050: the Potential for Demand-side Solutions to Greenhouse Gas Emissions Reduction. EuroChoices 2015, 14(3):4-11.
37. Revoredo-Giha C, Chalmers N, Akaichi F: Simulating the impact of carbon taxes on greenhouse gas emission and nutrition in the UK. Sustainability 2018, 10(1):134.
38. Säll S: Environmental food taxes and inequalities: Simulation of a meat tax in Sweden. Food Policy 2018, 74:147-153.
39. Slade P: The effects of pricing Canadian livestock emissions. Can J Agric Econ/Revue canadienne d'agroeconomie 2018, 66(2):305-329.
40. Springmann M, Mason-D'Croz D, Robinson S, Wiebe K, Godfray HCJ, Rayner M, Scarborough P: Mitigation potential and global health impacts from emissions pricing of food commodities. Nat Clim Chang 2016.
41. Springmann M, Sacks G, Ananthapavan J, Scarborough P: Carbon pricing of food in Australia: an analysis of the health, environmental and public finance impacts. Aust N Z J Public Health 2018, 42(6):523-529.
42. Vandenberghe D, Albrecht J: Tackling the chronic disease burden: are there co-benefits from climate policy measures? Eur J Health Econ 2018, 19(9):1259-1283.
43. Wirsenius S, Hedenus F, Mohlin K: Greenhouse gas taxes on animal food products: rationale, tax scheme and climate mitigation effects. Clim Change 2011, 108(1):159-184.
44. Zech KM, Schneider UA: Carbon leakage and limited efficiency of greenhouse gas taxes on food products. J Clean Prod 2019, 213:99-103.
