## Supplementary material for "Can a greenhouse gas emissions tax on food also be healthy and equitable? A systematized review and modelling study from Aotearoa New Zealand": Cheers checklist

### Consolidated Health Economic Evaluation Reporting Standards (CHEERS) 2022 Checklist

The CHEERS 2022 statement replaces the 2013 CHEERS statement, which should no longer be used. The CHEERS 2022 checklist contains 28 items with accompanying descriptions. Checklist users should indicate the section of the manuscript where relevant information can be found. The authors recommend using a section heading with a paragraph number. If an item does not apply to a particular economic evaluation, checklist users are encouraged to report "Not Applicable." If information is otherwise not reported, checklist users are encouraged to write, "Not Reported." Users should avoid the term "Not Conducted" as CHEERS is intended to guide and capture reporting. Additional information on CHEERS 2022 can be found here.

#### Title

##### 1. Title

Identify the study as an economic evaluation and specify the interventions being compared.

Page 1. Identified as a modelling study and states the focus of the work as greenhouse gas emissions taxes on food

#### Abstract

##### 2. Abstract

Provide a structured summary that highlights context, key methods, results, and alternative analyses.

Page 3.

#### Introduction

##### 3. Introduction: Background and Objectives

Give the context for the study, the study question, and its practical relevance for decision making in policy or practice.

Context of the study: Page 4, paragraph 4. Aim of study: Page 4, paragraph 4. Practical relevance: Page 4, paragraph 4.

#### Methods

##### 4. Health economic analysis plan

© 2022 ISPOR—The Professional Society for Health Economics and Outcomes Research
International Society for Pharmacoeconomics and Outcomes Research, Inc

Indicate whether a health economic analysis plan was developed and where available.

| |
|---|
| Not applicable |

##### 5. Study population

Describe characteristics of the study population (such as age range, demographics, socioeconomic, or clinical characteristics).

| |
|---|
| Page 5, paragraph 3 and Page 6, paragraph 2. |

##### 6. Setting and location

Provide relevant contextual information that may influence findings.

| |
|---|
| Not applicable |

##### 7. Comparators

Describe the interventions or strategies being compared and why chosen.

| |
|---|
| Page 6, paragraph 2 |

##### 8. Perspective

State the perspective(s) adopted by the study and why chosen.

| |
|---|
| Supplementary material, page 3, paragraph 6. |

##### 9. Time horizon

State the time horizon for the study and why appropriate.

| |
|---|
| Page 6, paragraph 2. |

##### 10. Discount rate

Report the discount rate(s) and reason chosen.

| |
|---|
| Supplementary material, page 3, paragraph 6. |

##### 11. Selection of outcomes

Describe what outcomes were used as the measure(s) of benefit(s) and harm(s).

| |
|---|
| Page 6, paragraph 2. |

##### 12. Measurement of outcomes

Describe how outcomes used to capture benefit(s) and harm(s) were measured.

| |
|---|
| Page 6, paragraph 2 and 4. Supplementary material, page 2, paragraph 5, page 3 paragraph 1 to 5 |


© 2022 ISPOR—The Professional Society for Health Economics and Outcomes Research
International Society for Pharmacoeconomics and Outcomes Research, Inc

##### 13. Valuation of outcomes

Describe the population and methods used to measure and value outcomes.

Supplementary material, page 2, paragraph 4-5, page 3 paragraph 1-6.

##### 14. Measurement and valuation of resources and costs

Describe how costs were valued.

Supplementary material, page 3, paragraph 5.

##### 15. Currency, price date, and conversion

~~Report the dates of the estimated resource quantities and unit costs, plus the currency and year of conversion.~~

Supplementary material, page 3, paragraph 5.

##### 16. Rationale and description of model

If modeling is used, describe in detail and why used. Report if the model is publicly available and where it can be accessed.

Supplementary material, page 2, paragraph 5, page 3 paragraph 1 to 5. Model is not publicily available.

##### 17. Analytics and assumptions

Describe any methods for analyzing or statistically transforming data, any extrapolation methods, and approaches for validating any model used.

not applicable

##### 18. Characterizing heterogeneity

Describe any methods used for estimating how the results of the study vary for subgroups.

Supplementary material, page 4, supplementary table 1

##### 19. Characterizing distributional effects

Describe how impacts are distributed across different individuals or adjustments made to reflect priority populations.

Page 7, paragraph 3

##### 20. Characterizing uncertainty

Describe methods to characterize any sources of uncertainty in the analysis.

Supplementary material, page 3, paragraph 6, page 4, supplementary table 1

~~21.~~

##### 21. Approach to engagement with patients and others affected by the study



© 2022 ISPOR—The Professional Society for Health Economics and Outcomes Research

International Society for Pharmacoeconomics and Outcomes Research, Inc

Describe any approaches to engage patients or service recipients, the general public, communities, or stakeholders (eg, clinicians or payers) in the design of the study.

| |
|---|
| Supplementary material, page 4, supplementary table 1. |

#### Results

##### 22. Study parameters

Report all analytic inputs (eg, values, ranges, references) including uncertainty or distributional assumptions.

| |
|---|
| Supplementary material, page 4, supplementary table 1. |

##### 23. Summary of main results

Report the mean values for the main categories of costs and outcomes of interest and summarize them in the most appropriate overall measure.

| |
|---|
| Page 13, table 2 |

##### 24. Effect of uncertainty

Describe how uncertainty about analytic judgments, inputs, or projections affects findings. Report the effect of choice of discount rate and time horizon, if applicable.

| |
|---|
| Uncertainty reported as uncertainty intervals throughout the paper e.g., page 13, table 2. Differing discount rates: page 14, table 3. time horizon. supplementary material. page 14. supplemental table 5 |

##### 25. Effect of engagement with patients and others affected by the study

Report on any difference patient/service recipient, general public, community, or stakeholder involvement made to the approach or findings of the study.

| |
|---|
| Not applicable |

#### Discussion

##### 26. Study findings, limitations, generalizability, and current knowledge

Report key findings, limitations, ethical, or equity considerations not captured and how these could impact patients, policy, or practice.

| |
|---|
| Key findings: page 15, paragraph 1 to 4, limitations: page 15, paragraph 5 to 6, page 16, paragraph 1 to 2, equity considerations: page 15. paragraph 3. policy impacts: page 16. paragraph 3 to 4. |


© 2022 ISPOR—The Professional Society for Health Economics and Outcomes Research
International Society for Pharmacoeconomics and Outcomes Research, Inc

#### Other Relevant Information

##### 27. Source of funding

Describe how the study was funded and any role of the funder in the identification, design, conduct, and reporting of the analysis.

Page 1, paragraph 1.

##### 28. Conflicts of interest

Report authors' conflicts of interest according to journal or International Committee of Medical Journal Editors requirements.

Page 2, paragraph 3.


© 2022 ISPOR—The Professional Society for Health Economics and Outcomes Research
International Society for Pharmacoeconomics and Outcomes Research, Inc
